# Aperiodic and oscillatory activity of the human brain during induced emotional states

**DOI:** 10.64898/2026.06.02.26354146

**Authors:** Haeorum Park, Carl Hacker, Hohyun Cho, Tao Xie, Alexis N. Simmons, Gansheng Tan, Eric C. Leuthardt, Peter Brunner, Jon T. Willie

## Abstract

Normal emotional experience depends on dynamic modulation of neural excitability across limbic and prefrontal circuits, yet the spectral markers that reflect these shifts in humans remain incompletely understood. In this study, we combined a validated video-based emotion induction paradigm with stereotactic electroencephalography (SEEG) in 31 patients with drug-resistant epilepsy to investigate how positive and negative affective states modulate oscillatory and aperiodic (asynchronous) neural activity. Using spectral parameterization to dissociate oscillatory power from the aperiodic 1/f component, we found that emotional valence robustly altered the aperiodic slope in a regionally specific manner: negative valence flattened the slope in thalamus, posterior insula, and posterior cingulate cortex, whereas positive valence produced flattening in dorsolateral prefrontal cortex. Simultaneous oscillatory changes included increased high-frequency activity and decreased alpha/beta power during negative affect, and reduced alpha power during positive affect, which were elucidated after adjusting for broadband aperiodic spectral shifts. These effects persisted after controlling for audiovisual stimulus or physiological features and were not evident in simultaneously recorded scalp EEG, underscoring their localization to intracranial sites. Together, these results provide the first direct evidence that active induction of emotional states modulates the aperiodic slope of human intracranial field potentials, reflecting valence-dependent shifts in local circuit excitability. The findings highlight the 1/f slope as a sensitive neural marker of affective brain states and for mood dysregulation.

**Graphical Abstract:** 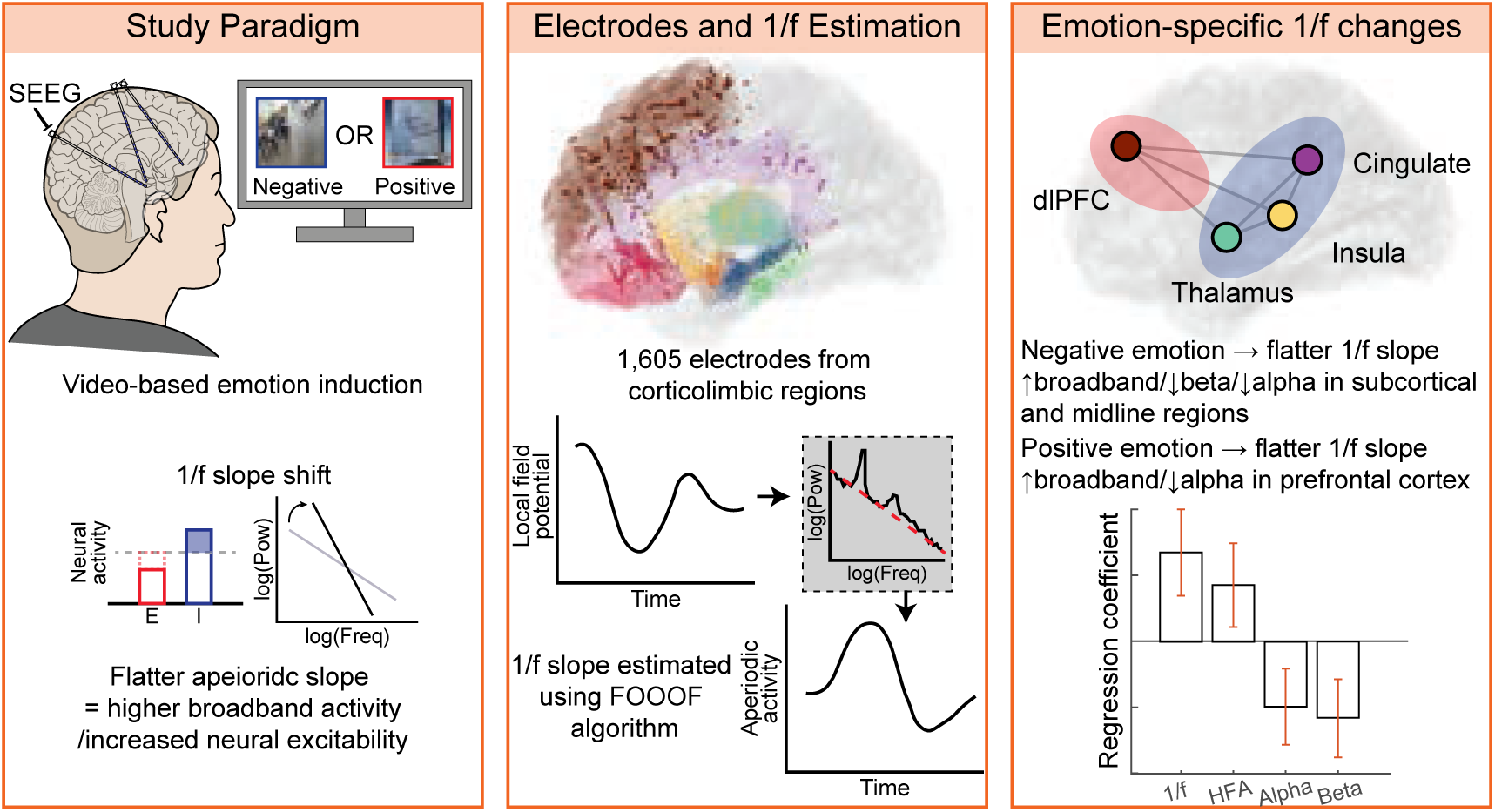

**Highlights:**

- Emotional valence modulates broadband neural activity: Using intracranial SEEG recordings, transient emotional states were shown to alter the aperiodic 1/f slope of human cortical and subcortical spectra in a regionally specific manner.
- Valence-specific circuit engagement: Negative emotion flattened the 1/f slope in the thalamus, posterior insula, and posterior cingulate cortex, whereas positive emotion induced similar flattening in the dorsolateral prefrontal cortex.
- Distinct oscillatory patterns accompany broadband shifts: Negative affect increased highfrequency activity and suppressed alpha/beta oscillation in subcortical and midline regions, while positive affect primarily reduced alpha power in the prefrontal cortex.
- 1/f slope as a marker of affective state: Findings identify the aperiodic spectral slope as a sensitive index of valence-dependent shifts in local neural excitability and a potential biomarker for mood-related dysregulation.

## 1. Introduction

Normal brain function depends on a delicate balance between excitatory and inhibitory (E/I) activity within neural circuits. In animal models, directly increasing or decreasing glutamatergic (excitatory) or GABAergic (inhibitory) signaling profoundly alters network dynamics and behavior (Sarawagi et al., 2021). In humans, E/I balance is thought to underlie various aspects of cognition and emotion, yet direct measurement of this balance remains challenging. Disruptions in E/I homeostasis have been implicated in several neuropsychiatric conditions – for example, enhanced cortical glutamate levels (reflecting a possible E/I imbalance) have been observed in depression, potentially contributing to symptoms such as negative affect and anhedonia (Haroon et al., 2018, Park et al., 2025). These findings suggest that mood and emotional processing may be strongly influenced by the excitation-to-inhibition ratio in key brain regions involved in affect regulation.

A major challenge in assessing E/I balance in the living human brain is the dependency on measuring indirect biomarkers. For example, traditional neuroimaging approaches, such as magnetic resonance spectroscopy, can estimate neurotransmitter levels or metabolic markers of neural activity, but cannot capture the moment-to-moment fluctuations of excitation versus inhibition (Haroon et al., 2018, Park et al., 2025). In contrast, noninvasive electrophysiological measures (EEG/MEG) provide insight into neural oscillations (e.g., alpha rhythms associated with inhibitory interneuron activity, gamma with excitation) that are often linked to inhibitory or excitatory processes (Lozano-Soldevilla et al., 2014). However, standard band-limited power analyses may not fully reflect the underlying E/I tone of a region, as they can be confounded by broad shifts in the power spectrum that are not tied to any single frequency band, but rather represent a change in the asynchronous *aperiodic* neural background activity (Miller et al., 2009). For example, changes in this aperiodic background (the overall 1/f-like spectral trend) can occur independently of narrow-band oscillations, potentially obscuring or mimicking changes in specific oscillatory power (Donoghue et al., 2020, Brake et al., 2024). This limitation has motivated the search for alternative spectral markers that capture global changes in network activity.

Recently, attention has turned to the aperiodic component of the neural power spectrum – the characteristic 1/f-like decrease of power with frequency – as a potential indicator of the brain’s global excitatory-inhibitory balance or related aspects of neural network excitability (Gao et al., 2017, Ahmad et al., 2022, Xiao et al., 2023, Cross et al., 2025). Unlike discrete oscillatory peaks, this aperiodic activity (quantified by the slope of the spectrum in log-log space) represents the “background” neural signal level (Brake et al., 2024). Computational modeling and empirical evidence suggest that modulating the balance of excitation and inhibition can lead to systematic changes in this spectral slope (**Figure 1**) (Gao et al., 2017). Intuitively, a flatter or less negative slope – indicating relatively higher power at high-frequency activity – has been hypothesized to reflect increased excitation or reduced inhibition. Conversely, a steeper or more negative slope – more power in low frequencies relative to high – may indicate a greater influence of inhibitory processes. **Figure 1** illustrates this concept: in a network with relatively more excitatory drive (or less inhibition), high-frequency activity is enhanced and the 1/f slope flattens, while a network dominated by inhibition shows a pronounced 1/f drop-off (steeper slope). Consistent with this idea, some studies have reported that pharmacologically reducing cortical E/I ratios (for instance, using a GABAergic anesthetic in primates) produces a noticeably steeper EEG power spectrum, tracking loss of consciousness (Gao et al., 2017, Pertermann et al., 2019). Conversely, states of heightened neural activation tend to show elevated high-frequency power and a shallower spectral slope, a pattern that has been interpreted as reflecting increased “neural noise” from asynchronous spiking (Voytek et al., 2015). If the 1/f slope indeed indexes aspects of cortical excitability, it could provide a valuable, noninvasive window into neural circuit function that complements traditional oscillation measures.

**Figure 1.**
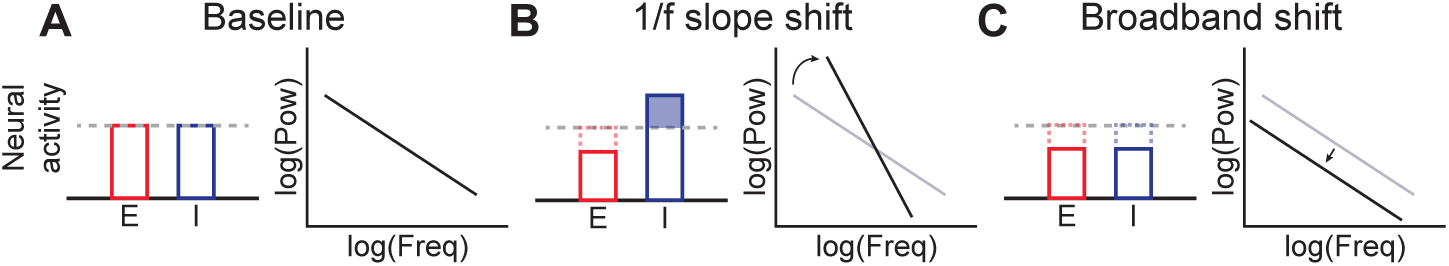
Schematic of excitatory and inhibitory activity changes and aperiodic spectral features. **(A)** Example neural power spectrum with a typical excitatory and inhibitory activity. **(B)** Decreased E/I balance (i.e., decreased excitatory and/or increased inhibitory neural activity) reflected as decreased (i.e., steeper) 1/f slope. **(C)** Decreased overall neural activity, including excitatory and inhibitory, reflected as decreased high-frequency broadband activity.

Nevertheless, it is important to emphasize that the mechanistic interpretation of the aperiodic slope remains an open question. Growing evidence indicates that the 1/f spectral exponent is shaped by a variety of biophysical and network phenomena, rather than serving as a specific readout of E/I balance alone (Brake et al., 2024). Many different mechanisms have been proposed to underlie 1/f-like scaling in neural power spectra including (i) dendritic and circuit-level low-pass filtering, which attenuates high-frequency currents more strongly and thereby generates a 1/f-like decay (Buzsáki et al., 2012), (ii) frequency-dependent conductivity and permittivity of brain tissue leading to filtering effects through ionic diffusion and polarization (Bédard and Destexhe, 2009), (iii) the superposition of many damped oscillatory processes with a distribution of relaxation rates, which can parsimoniously explain both alpha blocking and 1/f scaling in resting EEG (Evertz et al., 2022), and (iv) synaptic timescale-based explanation, in which asynchronous inputs filtered by AMPA and GABA receptor kinetics give rise to Lorentzian-like spectral trends that shape the background EEG component (Brake et al., 2024). Together, these perspectives suggest that the 1/f slope is unlikely to serve as a single, specific marker of E/I balance, but rather reflects a composite background signal shaped by dendritic filtering, tissue properties, damped population oscillations, and synaptic kinetics.

Furthermore, the aperiodic exponent can be viewed more generally as an index of neuronal noise or population firing variability. In this view, a flatter spectrum corresponds to more unsynchronized, random firing (higher “noise”), whereas a steeper spectrum implies more correlated, synchronous firing (lower noise) (Voytek et al., 2015). Consistent with this, in silico work has shown that simply introducing stochastic fluctuations into network models can produce 1/f-like spectral scaling without requiring any specific change in E/I balance or critical-state dynamics (Kramer and Chu, 2024). Recent experimental studies also urge caution in equating the aperiodic slope with excitation/inhibition levels: for instance, one rodent EEG study found that manipulating the E/I ratio did not lead to predictable changes in the spectral exponent, calling into question the reliability of 1/f slope as a marker of E/I in isolation (Salvatore et al., 2024). In summary, while changes in the 1/f slope do reflect alterations in underlying neural activity, this measure likely captures a broad composite of network factors (e.g., overall synaptic input, firing irregularity, and other “background” processes) rather than serving as a precise readout of the E/I ratio alone (Brake et al., 2024). We therefore interpret the aperiodic slope in a nuanced way – as a general indicator of the brain’s state of network excitability or arrhythmic activity (sometimes termed “background noise”) – and avoid making overly specific claims that it directly indexes E/I balance in our analyses.

Given the above, there is still strong motivation to study the 1/f slope as a complementary feature of neural activity, especially in contexts where broad shifts in network state are expected. Differences in the aperiodic exponent have been linked to a variety of functional and clinical variables – including age, cognitive performance, arousal state (anesthesia vs. wake), sleep stages, and neurological disorders (Voytek et al., 2015, Pertermann et al., 2019, Ostlund et al., 2021, Brake et al., 2024) – suggesting that this metric is sensitive to meaningful changes in brain function. Applying this concept to affective neuroscience, one can hypothesize that transient emotional states involve measurable modulations of circuit-level excitability or inhibitory tone in brain regions important for mood regulation. Positive and negative emotions engage distributed networks that include limbic and prefrontal areas (such as the anterior cingulate cortex, orbitofrontal cortex, and insula), which integrate sensory information with emotional valence and internal state (Kim et al., 2025). It is plausible that transitions into different emotional states could shift local neural dynamics in these regions – for example, a negative or sad experience might tilt circuits toward heightened inhibitory reactivity. In clinical depression, an inability to appropriately modulate these dynamics (e.g., failure to engage inhibitory calming mechanisms, or hypoactive baseline noise) might manifest as blunted positive affect or exaggerated negative emotional responses. However, directly testing these ideas in humans is difficult, given the lack of any noninvasive methods to track E/I fluctuations in real time during emotional processing.

Stereotactic electroencephalography (SEEG) provides a rare opportunity to investigate the source of human neural dynamics, including within deep limbic circuits, with high temporal resolution. In patients undergoing SEEG for epilepsy monitoring, depth electrodes are stereotactically inserted into various brain regions, often sampling structures implicated in emotion. While these recordings primarily serve clinical seizure localization, they offer a unique window into in vivo human neurophysiology. In the present study, we leveraged SEEG in a large cohort of epilepsy patients to examine how induced emotional states affect the spectral features of local field potential – in particular, the aperiodic 1/f slope, a candidate measure of local network excitability (sometimes linked to E/I balance). We employed a video-based emotion induction paradigm using validated emotional clips (Samide et al., 2020) to evoke positive, negative, and neutral affective states while recording SEEG from regions including the anterior cingulate cortex (ACC) and orbitofrontal cortex (OFC), which are crucial for emotional experience and regulation. Simultaneous scalp EEG was also recorded to assess whether spectral effects of induced emotion were apparent at the cortical surface or were localized to intracranial sites.

We hypothesized that transitions between emotional states would modulate both traditional oscillatory power and the aperiodic component of the SEEG power spectrum in limbic circuits. Specifically, we anticipated that emotionally valenced stimuli might produce broadband changes in power and corresponding shifts in the 1/f slope, reflecting changes in underlying neural excitability. To dissociate narrow-band oscillatory changes from global spectral shifts, we applied a spectral parameterization approach, the FOOOF algorithm (Fitting Oscillations and One-Over-F), to each power spectrum (Donoghue et al., 2020), fitting the aperiodic exponent over the 2-50 Hz range while accounting for any superimposed oscillatory peaks. The selected frequency range was determined based on two considerations: first, to encompass all canonical narrow-band oscillations (delta, theta, alpha, beta, and gamma), thereby capturing the global spectral structure; and second, because prior work has demonstrated a relationship between the E/I ratio and the 1/f slope, particularly within the 30-50 Hz range (Donoghue et al., 2020, Hacker et al., 2025, Gao et al., 2017). By comparing spectral measures, before and after removing the aperiodic “background”, we aimed to determine whether emotion-related effects were driven by specific band-limited oscillations or by broadband shifts in the spectrum. Additionally, we explored whether any such effects were detectable on simultaneously recorded scalp EEG (which would indicate more diffuse cortical involvement) or were unique to intracranial recordings. This study thus integrates a novel analytic framework (spectral parameterization of aperiodic activity) with a unique intracranial dataset to probe the neural correlates of emotion. Our findings shed light on how changes in emotional state impact both oscillatory and aperiodic brain activity, and we discuss their implications for models of neural circuit function in affective processes (including the role of excitatory/inhibitory dynamics) and for potential biomarkers of mood dysregulation in disorders such as depression.

## 2. Methods

### Participants

A total of 31 drug-resistant epilepsy patients (age 18-69, 35±12 years, 17 males, 14 females) undergoing intracranial seizure monitoring as a part of their clinical care at the Barnes-Jewish Hospital or St. Louis Children’s Hospital participated in this study (demographic details in **Table S1**). All patients had normal intellectual function (IQ range 71-120, average 100) and gave informed consent to an IRB-approved research protocol in addition to their clinical monitoring. Depth electrodes (8-16 contacts each) were stereotactically implanted targeting potential seizure foci, as well as additional limbic, paralimbic, and frontal regions for comprehensive coverage. None of the participants was in an active seizure state during the recordings analyzed, nor had they experienced a seizure during the experiment. Any data epochs exhibiting interictal epileptiform activity were excluded from our analysis.

### Electrode localization

Each participant underwent a preoperative T1-weighted magnetic resonance imaging (MRI) scan to acquire structural brain images. Following intracranial electrode implantation, the participant underwent postoperative volumetric computed tomography (CT) scans. These postoperative CT images, in conjunction with three-dimensional subject-specific cortical models, are used to derive the electrode locations (Coon et al., 2016). For visualization purposes, individual data were warped to a common anatomical space (MNI152).

The implanted SEEG electrodes were approved for human use (Ad-Tech Medical Corp., Racine, WI; PMT Corp., Chanhassen, MN; DIXI Marchaux-Chaudefontaine, France). Trajectories were planned using a stereotactic software (ROSA, Zimmer Biomet, Inc., Warsaw, IN). Under general anesthesia and using sterile technique, 2.4 mm diameter twist-holes were drilled, and bolts were secured into the bone under robotic assistance to realize the desired path for each trajectory. The dura was penetrated, and depth electrodes were inserted to the desired depth and secured in each bolt. The platinum-iridium electrode contacts were 2 mm in length (0.8 mm in diameter) and spaced 3.5-5 mm center-to-center. Electrode contacts were localized by co-registering the preoperative T1-weighted MRI to a postoperative CT scan using Versatile Electrode Localization Framework (VERA) (Adamek et al., 2022) software (**Figure 2**). Post-implantation electrodes were connected to a Nihon Kohden JE-120A amplifier for continuous monitoring. Pial surface reconstructions were created from preoperative T1 MRI scans using Freesurfer (Fischl, 2012).

**Figure 2.**
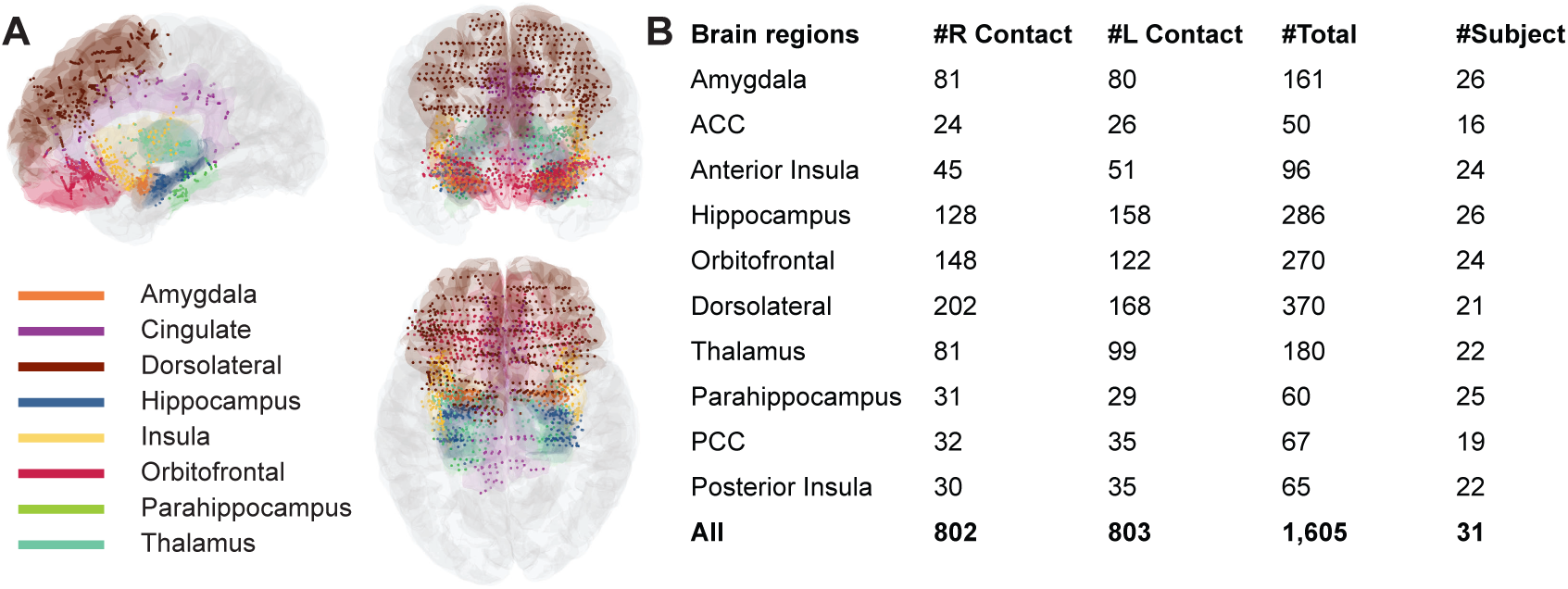
Electrodes placements. **(A)** 3D sliced view (sagittal, coronal, and transverse) of electrode locations for all subjects superimposed onto the reconstructed brain in Montreal Neurological Institute (MNI) coordinate space. **(B)** The number of electrode contacts in each region of interest (ROI) is noted in the table. We had a total of eleven ROIs with a total of 1,605 electrodes implanted. Each subject had partial ROI coverage since the depth electrode planning was part of clinical care.

We determined the regions of interest for our mood-related analyses based on a review paper on mood disorder-related networks (Williams, 2016). For instance, the posterior cingulate cortex was selected as it is part of the default mode network, and its activity and connectivity increase during depressive rumination (Greicius et al., 2003, Hamilton et al., 2015, Kim et al., 2025). All patients had one or more electrodes implanted in emotion-related cortico-limbic regions, containing structures implicated in emotion (Williams, 2016) (**Figure 2**). We sought to identify neurophysiological correlates of self-reported changes in emotion in these a priori regions. We examined 10 regions of interest: amygdala, anterior cingulate cortex (ACC), anterior insula, hippocampus, orbitofrontal cortex, dorsolateral cortex, temporal pole, thalamus, posterior cingulate cortex (PCC), and posterior insula. In the main analyses, the thalamus was treated as a single region; however, given the functional heterogeneity of thalamic nuclei, we additionally reported the distribution of thalamic contacts across nuclei (**Table S2**).

### Emotion induction task

The task in our study used a previously validated video database to investigate the dynamics of induced emotion (Samide et al., 2020). Patients watched a series of news clips, edited to 20-50 seconds in length, with a narrator explaining the situation in a clear and understandable way (**Figure 3**). The news clips covered a variety of scenarios, from emotional coverage of terrorism or war to situations expected to evoke positive emotions, such as celebrity weddings or donations. The emotional valence was uniformly distributed, ranged from positive to negative, and also included neutral clips (**Figure 3B**). Clips were presented without any interruptions in the patient’s private epilepsy monitoring unit (EMU) room using a monitor placed approximately 70 cm in front of the patient. Audio was delivered through speakers at a consistent volume. Participants were instructed to attend to each video and allow themselves to feel whatever emotion it prompted. After watching each video, they are asked to rate their current feeling on a Likert scale from 1 (very unpleasant) to 9 (very pleasant). A total of 40 video clips were shown. To avoid carry-over effects, a short break was taken after completing 20 trials. The final 10 seconds of each trial were used for spectral analysis to ensure that emotional change reached a saturated, stable level. We excluded trials with no or irrelevant responses. If a response on a particular trial exceeded two group standard deviations (e.g., a positive response to a news report about a shooting), it was excluded from further analysis. The self-reported valence ratings confirmed that the chosen clips successfully evoked the intended emotion (see Results).

**Figure 3.**
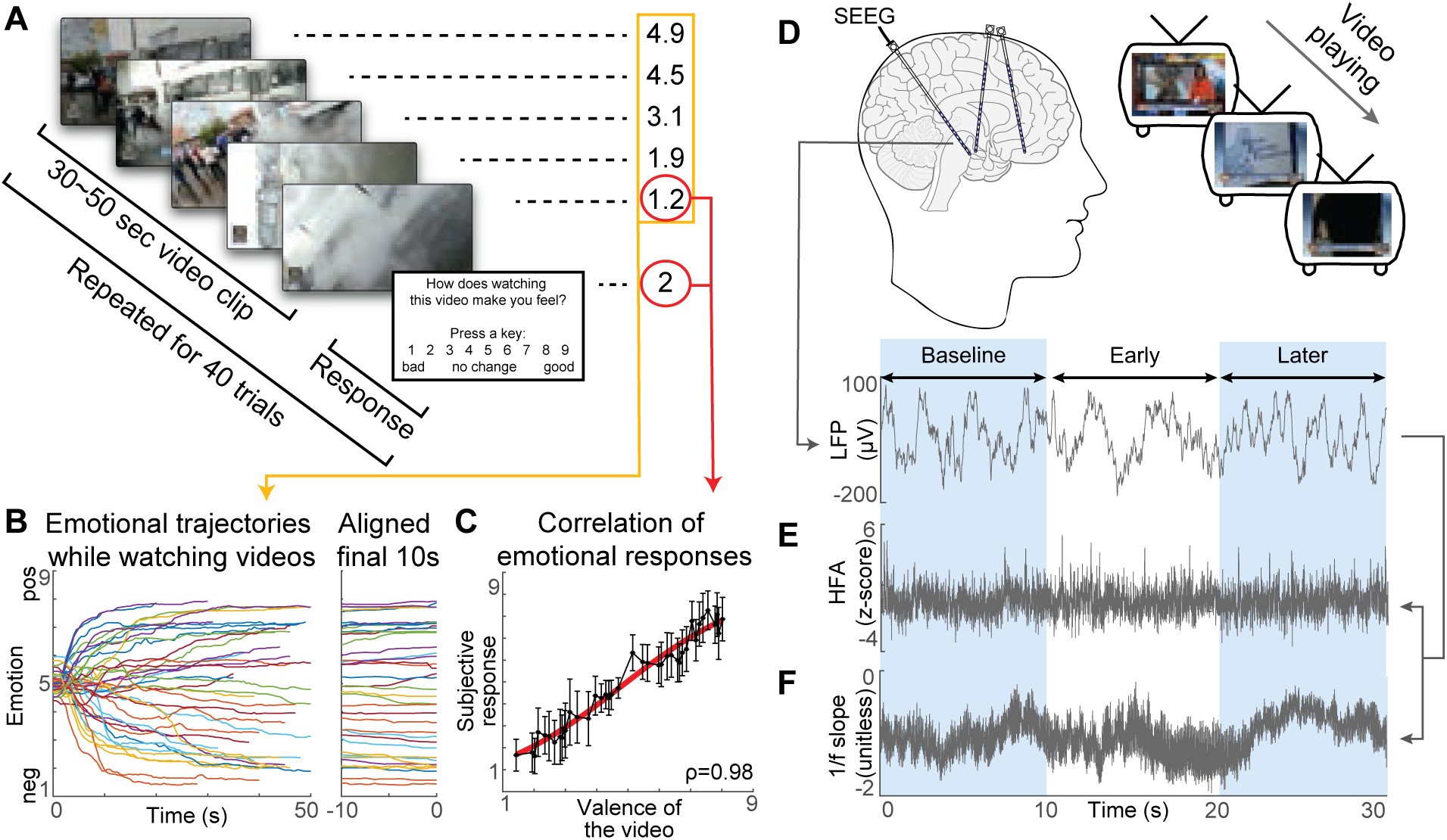
Emotion induction task and intracranial recording paradigm. **(A)** The newscaster explains the event depicted in each video clip (e.g., a geyser erupts under the bus). Emotional responses while watching the clip rated continuously by 100 participants, were included in the published dataset (orange rectangle) (Samide et al., 2020). In our study, participants reported their emotional states after viewing the entire clip (red circle). **(B)** Emotional dynamics of each video from the published dataset are shown (left). To assess stabilization, we aligned the final 10 seconds across videos (right), revealing that emotional responses were saturated and stable at the end of viewing. **(C)** Emotional ratings from our participants after viewing the clips showed a strong linear correlation with those from the published dataset, validating that our paradigm successfully induced the intended emotional states. **(D)** Epilepsy patients implanted with depth electrodes (SEEG) viewed the emotional video clips during intracranial recordings. Local field potentials (LFPs) were obtained from electrodes targeting multiple brain regions. The first 10 seconds of the LFP was used as the baseline and the later 10 seconds of the LFP was compared to the baseline. **(E)** Example preprocessing of LFP data to extract high-frequency activity (HFA). **(F)** Example preprocessing of LFP data to estimate the 1/f slope from the power spectrum.

### Intracranial recordings

We recorded SEEG signals from the subject at their bedside (**Figure 3A**) using the general-purpose Brain-Computer Interface (BCI2000) software (Schalk et al., 2004), interfaced with Nihon Kohden JE-120A long-term recording system (Nihon Kohden, Tokyo, Japan) to amplify, digitize, and store the signal (sampling rate 2,000 Hz). The recordings were obtained during quiet wakefulness.

Trials affected by interictal epileptiform discharges (iEDs) were detected using an automated iED detection algorithm (AiED) (Quon et al., 2022), and removed from further analysis. Local field potentials were recorded, and 60 Hz line noise power was calculated from each electrode. The signals were re-referenced to a common average calculated across those channels that exhibited non-significant line noise (i.e., three standard deviations). The local field potentials were then notch-filtered at 60 Hz and their harmonics (120 and 180 Hz) to reduce line noise-related artifacts, and the signals were downsampled to 500 Hz to simplify further analysis. (Cheung et al., 2016).

### Spectral analysis: Aperiodic and periodic components

We computed the power spectral density (PSD) of the SEEG signal using Welch’s method across the full clip duration for each electrode contact and video trial. Frequency bins at 60 Hz (and its harmonics) were removed to avoid line noise artifacts. This produced an average power spectrum for each contact in each condition (negative, neutral, positive). We quantified bandlimited power by averaging the PSD across the following frequency ranges: alpha (8-12 Hz), beta (12-30 Hz), and high-frequency activity (70-150 Hz). Subsequently, this activity was binned into 10-second epochs to identify changes induced by the presented news clips (**Figure 3E**).

To quantify the aperiodic 1/f component of the spectrum, we applied the FOOOF (Fitting Oscillations and One-Over-F) algorithm. FOOOF models the PSD as the sum of a power-law background (characterized by an exponent and offset) and any superimposed oscillatory peaks. We fit the model in the 2-50 Hz range for each contact’s spectrum, allowing the algorithm to capture the aperiodic slope while identifying any prominent oscillatory peaks. The selected frequency range was determined based on two considerations: first, to encompass all canonical narrow-band oscillations (delta, theta, alpha, beta, and gamma), thereby capturing the global spectral structure; and second, because prior work has demonstrated a relationship between the E/I ratio and the 1/f slope, particularly within the 30-50 Hz range. The resulting aperiodic exponent was taken as our measure of interest (**Figure 3F**). A lower (steeper) slope indicates relatively more low-frequency and less high-frequency power. We also computed 1/f-adjusted power spectra by subtracting the fitted aperiodic exponent component from the PSD, yielding residual periodic spectra.

### Statistical analysis

We used linear mixed-effects models (LMMs) to assess the effect of emotion valence on neural measures, while accounting for repeated measurements within participants and across brain regions. The primary analysis was structured to test whether emotional valence modulated the spectral features (aperiodic exponent and band power) relative to each trial’s baseline (**Figure 4A**). To this end, for each trial, we computed the change in each measure from baseline to the emotion epoch (later minus baseline). This change score captures the neural response induced by the video, controlling for each early-trial’s baseline. These change scores were then used as the dependent variable in the LMMs.

**Figure 4.**
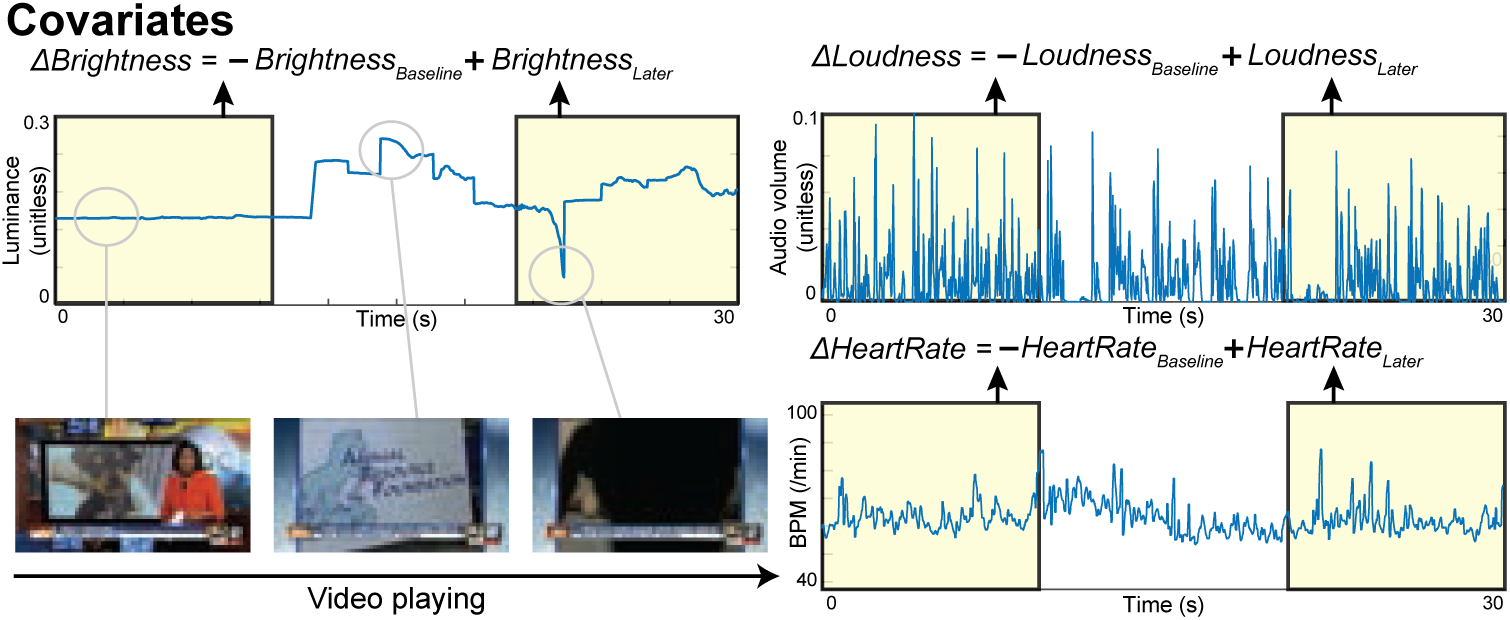
Statistical modeling framework for neural feature analyses. **(A)** Linear mixed-effects model (LMM) used for primary analyses: *Neural f eature* ∼ *Valence* + *Region* + *ValenceRegion* + (1|*S ub ject*) + (1|*S ub ject* : *Region*) + (1|*S ub ject* : *Region* : *Electrode*). This model tested whether emotional valence modulated neural spectral measures while accounting for subject-, region-, and electrode-level variability. **(B)** Secondary LMM including heart rate/audiovisual covariates to ensure robustness against low-level stimulus features: *Neural f eature* ∼ *Valence* + *Region* + *ValenceRegion* + Δ*HeartRate* + Δ*Loudness* + Δ*Brightness* + (1|*S ub ject*) + (1|*S ub ject* : *Region*) + (1|*S ub ject* : *Region* : *Electrode*). This model confirmed that effects of emotional valence were not explained by clip brightness or loudness. **(C)** Example time traces of brightness and loudness extracted from the video stimuli, illustrating how low-level audiovisual features varied across different segments (e.g., bright vs. dark scenes, speech vs. silence).

We first performed a behavioral check: a paired *t*-test compared the self-reported valence ratings between negative and positive clip conditions, and a Pearson’s correlation between the self-reported ratings and the emotional score provided from the database to confirm successful induction of different emotional states.

For neural data, separate LMMs were fit for each spectral measure (aperiodic exponent, raw alpha power, raw beta, raw HFA, adjusted alpha, adjusted beta). In each model, *Valence* was the main predictor. We also included *Brain Region* as a fixed factor to account for regional differences, and the interaction term *Valence* x *Region* to identify region-specific valence effects (**Figure 4A**). *Subject*, *Subject* x *Region*, and *Subject* x *Region* x *Electrode* were included as a random intercept to model individual differences. This approach effectively performs a within-subject comparison of the neural response to emotional stimuli, aggregated by region. Degrees of freedom and *p*-values for fixed effects were obtained via Satterthwaite approximation (Satterthwaite, 1946). Post-hoc contrasts (Tukey-corrected) were used to probe significant interactions, i.e., to determine regions that exhibit significant differences between valence and neutral. We performed two sets of analyses: one focusing on *Negative* vs. *Neutral* changes and the other on *Positive* vs. *Neutral*, for each region, in order to detail how each emotional valence condition differed from neutral.

To ensure robustness, we included several covariates in control analyses (**Figure 4B-C**). We considered heart rate variability (e.g., as a proxy of arousal, *HeartRate*) and low-level audiovisual features of the clips (e.g., mean luminance, *Brightness*; and mean sound volume, *Loudness*) as covariates in a secondary model, to check that any observed spectral changes were not merely due to differences in arousal or how bright or loud the stimuli were (no significant effect was found). Statistical analyses were carried out in MATLAB R2023a. We used the *fitlme* package in MATLAB for mixed effect modeling and *multcompare* for post-hoc tests. Significance was defined at *p* < 0.05 (two-tailed). When multiple comparisons were performed (e.g., testing multiple regions or bands), we applied family-wise error rate corrections (FWER) as appropriate, although in many cases the mixed model inherently handles multiple data points. All key results were also verified using nonparametric approaches, yielding consistent findings. Data visualization was done in MATLAB and refined in Adobe Illustrator. Results are reported with an emphasis on effect sizes (e.g., fixed-effect coefficients) alongside *p*-values to convey valence-driven changes.

## 3. Results

### Electrodes placements in emotion-related cortico-limbic regions

A total of 31 drug-resistant epilepsy patients participated in this study. All patients were implanted with intracranial electrodes solely to identify the localizing seizure. We had total of 1,605 electrodes located in emotion-related cortico-limbic region across 31 patients (**Figure 2B**), including electrodes in the amygdala (161 contacts from 26 subjects), cingulate cortex (117 contacts from 20 subjects), insular cortex (161 contacts from 26 subjects), thalamus (180 contacts from 22 subjects), and dorsolateral prefrontal cortex (dlPFC, 370 contacts from 24 subjects) etc.

### Behavioral validation of emotional induction

We first confirmed that the video stimuli used in this study reliably modulated participants’ reported affective state. The video database (Samide et al., 2020) provided continuous self-report emotion traces recorded while each participant watched the clips (**Figure 3A**). Examination of these traces revealed that emotions began in a neutral range at clip onset, progressively diverged during the middle portion, and stabilized during the final 10 s (**Figure 3B**). Accordingly, we defined the last 10 s as the *later* (emotion-induced) phase and the first 10 s as the *baseline* (relatively neutral) phase for all subsequent analyses (**Figure 3D-F**).

Participants’ self-report ratings confirmed that the video clips successfully induced the targeted emotional valences. On a scale from 1 (very negative) to 9 (very positive), negative clips were rated as strongly unpleasant (mean valence ≈ 2.1 ± 0.64), whereas positive clips were rated as pleasant (mean valence ≈ 7.6 ± 0.52). The difference in valence ratings between negative and positive conditions was highly significant (*t_d f_* > 10, *p* < 0.0001), indicating a clear separation of emotional experience. Moreover, individual ratings showed a robust linear correspondence with the normative valence scores reported in the original video database (*p* < 0.0001; **Figure 3C**) (Samide et al., 2020). These behavioral results establish the validity of our emotion-induction approach, providing a foundation for interpreting the neural changes associated with negative and positive affective states.

### Aperiodic 1/f slope changes with emotional valence

We observed significant modulations of the aperiodic spectral exponent (1/f slope) in subcortical and prefrontal circuits as a function of emotional valence. **Figure 3F** illustrates a representative aperiodic exponent time trace from a contact in the thalamus during a negative video and its preceding baseline. The negative emotion epoch exhibits a noticeably flatter 1/f slope (i.e., less negative exponent). Across participants, negative valence consistently induced a flatter spectral slope (increased aperiodic exponent χ) in several subcortical and midline regions. In particular, the thalamus, posterior insula (pIns), and posterior cingulate cortex (PCC) exhibit significant exponent increases during negative videos compared to baseline (LMM Valence effect for *Negative* vs. *Baseline* in these regions, *p_FWER_* < 0.001, corrected) (**Figure 5A-D,I**). Because of the functional heterogeneity of the thalamic nuclei, we replicated the analyses by discriminating among the nuclei (**Figure S1**). Please see **Table S2** and **Figure S1** for details. The mean exponent elevation in these areas ranged from Δχ ≈ 0.3 to 0.5 (unitless, representing the change in slope of the log-log PSD). No other regions exhibited a significant decrease in slope for negatively-valenced stimuli; in these areas, the effect was a selective flattening of the aperiodic component, suggestive of increased high-frequency activity.

**Figure 5.**
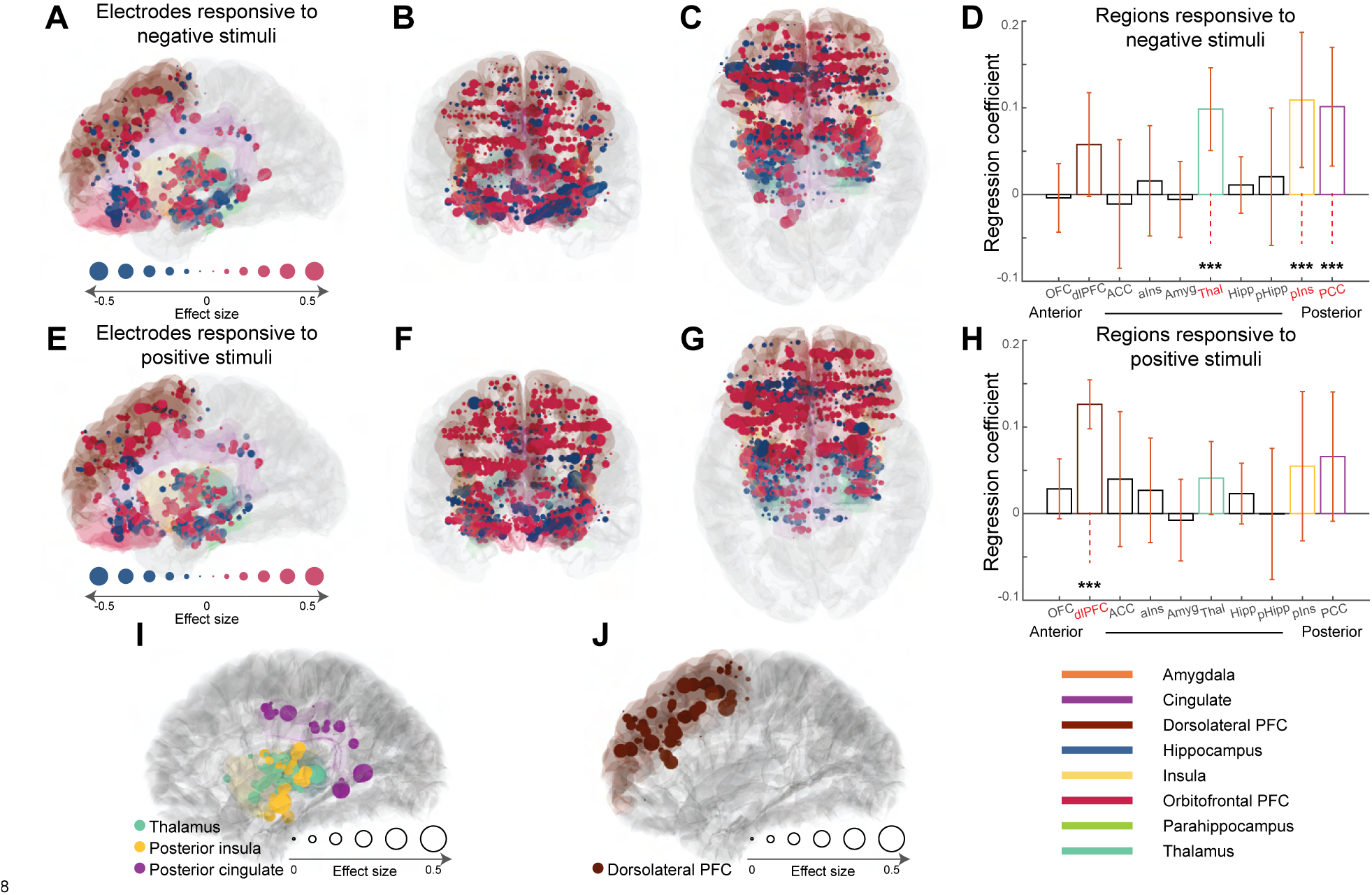
Brain regions and electrode responses showing valence-related changes in the aperiodic exponent (1/f slope). **(A-C)** Sagittal, coronal, and transverse views showing electrode contacts whose aperiodic exponent changed in response to negative video stimuli. Circle size reflects the Pearson’s correlation coefficient between emotional valence and the 1/f slope. Larger red circles indicate electrodes where the 1/f slope became flatter during more negative stimuli, whereas blue circles indicate electrodes where the slope became steeper. **(D)** Linear mixed-effects model (LMM) results summarizing regional effects for negative valence. Bars represent regression coefficients for each region of interest. Significant effects were observed in the thalamus, posterior cingulate cortex (PCC), and posterior insula (pIns). Error bars (orange) indicate 99.5% confidence intervals. **(E-G)** Sagittal, coronal, and transverse views showing electrode responses to positive video stimuli, using the same visualization scheme as in A-C. **(H)** LMM results summarizing regional effects for positive valence. A significant regression coefficient was observed in the dorsolateral prefrontal cortex (dlPFC). **(I)** Subset of electrodes in the thalamus, pIns, and PCC that were responsive to negative valence, demonstrating the consistency of effects within these regions. **(J)** Subset of electrodes in the dlPFC that were responsive to positive valence. Circle size reflects the Pearson’s correlation coefficient between emotional valence and the 1/f slope. Significance thresholds: *** *p_FWER_* < 0.0001.

In contrast, positive valence produced a notable flattening of the 1/f slope predominantly in the dorsolateral prefrontal cortex (dlPFC) (**Figure 5E-H,J**). During positive emotion epochs, dlPFC contacts showed a higher exponent (flatter slope) compared to baseline (mean Δχ ≈ 0.2), a change that was significant in our correlation analysis (*p_FWER_* < 0.001). Other regions, including the amygdala, anterior cingulate, anterior insula, orbitofrontal cortex, and hippocampus, did not show consistent valence-driven changes in the exponent.

Directly comparing the two valence conditions, we found a significant *Valence* x *Region* interaction for aperiodic exponent (*p* < 0.01). Post-hoc contrasts revealed that for negative vs. positive valence, the thalamus, pIns, and PCC each exhibit a significantly flatter slope for negative valence (i.e., χ_negative_ > χ_positive_, *p* < 0.01). In contrast, the dlPFC exhibits the opposite pattern (flatter in positive than in negative condition, *p* < 0.05). In other words, negative emotion preferentially impacted the 1/f slope in subcortical and midline regions – thalamus, pIns, and PCC – whereas positive emotion affected the prefrontal area. **Figure 5D, H** plots the group-mean of the aperiodic exponent during baseline and emotion-conditioned periods for these key regions, highlighting the divergent effects of valence.

It is worth noting that not all regions within the emotion-related cortico-limbic network responded uniformly: for example, the amygdala did not exhibit a reliable slope change with negative valence in our data, a significant finding that is addressed in the *Discussion*. Nonetheless, these findings provide clear evidence that emotional states can transiently shift the aperiodic 1/f slope in human brain field potentials, in a regionally specific manner tied to valence.

### Oscillatory power changed in alpha, beta, and high frequencies

Along with changes in the aperiodic component, we found significant emotion-related modulations in traditional oscillatory frequency bands. These effects were apparent in 1/f adjusted power spectra (i.e., power after removing the 1/f fit) (**Figure 6**). For negative emotions, multiple subcortical and midline regions exhibited decreases in low-frequency oscillatory power alongside increases in high-frequency activity (HFA), consistent with an overall activation pattern. Specifically, during negative emotion the thalamus showed a significant increase in HFA (70-150 Hz) power relative to baseline (mean increase +0.5 dB, *p_FWER_* < 0.01) and a concurrent decrease in alpha (8-12 Hz) power (mean decrease −0.4 dB, *p_FWER_* < 0.05) (**Figure 6A-C**). This indicates that thalamic activity shifted from low-frequency fluctuations to high-frequency fluctuations when processing negative stimuli. Similarly, the posterior insula (pIns) exhibited a suppression of alpha and beta power in the negative condition: pIns alpha dropped by −0.3 (dB) and beta (12-30 Hz) by −0.2 (dB), both significant compared to baseline (*p_FWER_* < 0.01). The posterior cingulate cortex (PCC) showed a reduction in beta power during negative emotion (−0.3 dB, *p_FWER_* < 0.05). No significant HFA increase was detected in PCC for negative clips, but there was a trend toward higher HFA in pIns.

**Figure 6.**
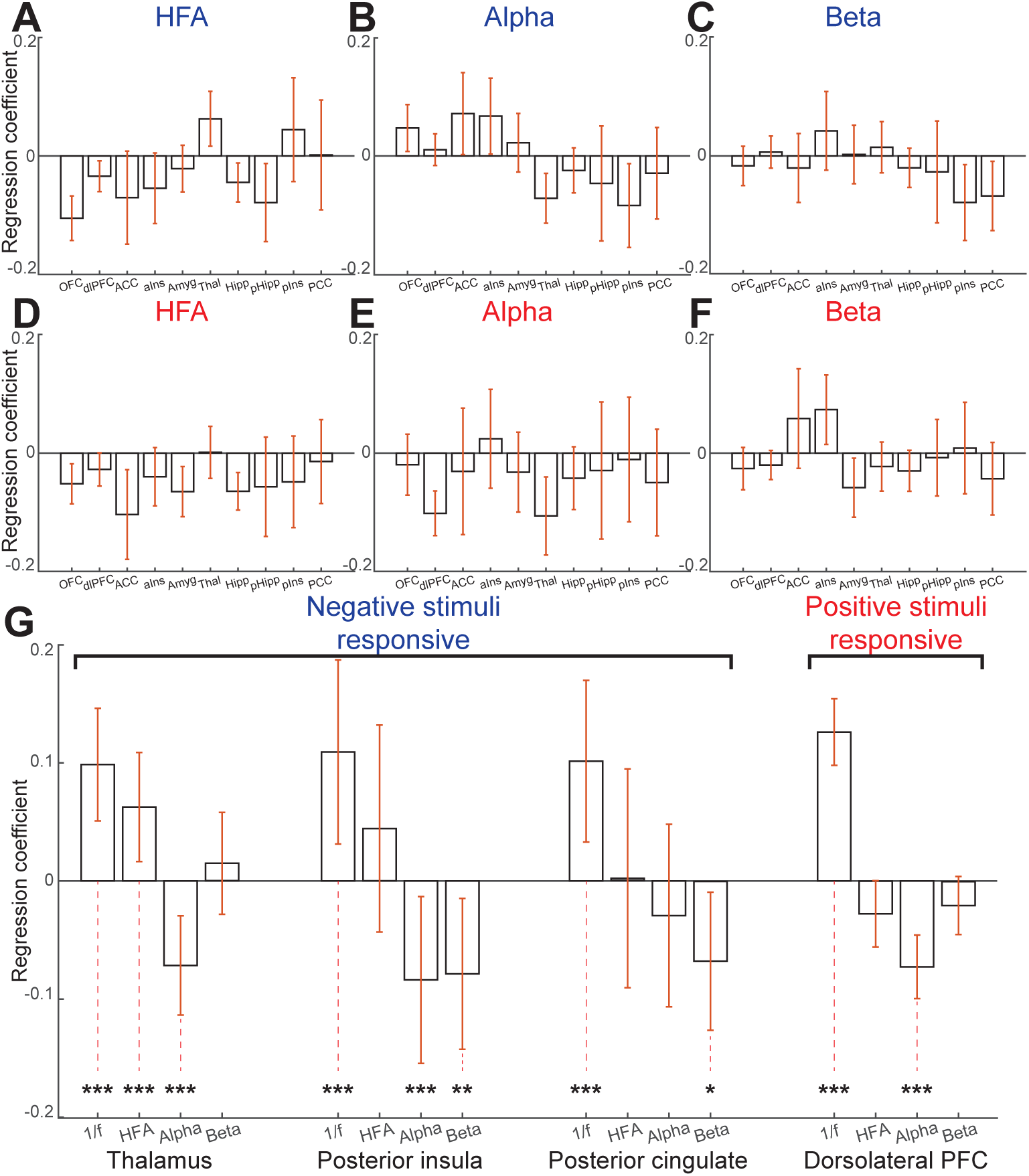
Periodic power changes in slope-responsive ROIs after 1/f adjustment. **(A–C)** Detrended high-frequency activity (HFA), alpha, and beta power during viewing of negative emotional videos. **(D–F)** Detrended HFA, alpha, and beta power during viewing of positive emotional videos. **(G)** Summary of periodic power changes across slope-responsive ROIs. Alpha and beta power are shown after 1/f correction. Error bars (orange) indicate the 99.5% confidence interval. Significance thresholds: *** *p_FWER_* < 0.001, ** *p_FWER_* < 0.01, * *p_FWER_* < 0.05.

In contrast, positive emotion primarily affected the dlPFC in terms of oscillatory power; the dlPFC showed a significant decrease in alpha power during positive videos compared to baseline (mean −0.3 dB, *p_FWER_* < 0.001) (**Figure 6D-F**). This alpha suppression in dlPFC suggests increased engagement or excitation of this region during positive affect, as alpha is inversely related to cortical activation (Bönstrup et al., 2015). We did not observe a significant HFA increase in dlPFC for positive emotion at the group level. For other regions, the anterior insula showed increased beta oscillatory power for both positive and negative video, and the orbitofrontal cortex and hippocampus HFA were decreased for both positive and negative video stimuli.

To summarize the oscillatory power findings: Negative valence (vs. baseline) was associated with increased high-frequency power and decreased alpha/beta power in key subcortical regions (thalamus, pIns, PCC), and positive valence (vs. baseline) was associated with decreased alpha power in dlPFC (**Figure 6G**). These patterns align with an overall schema that negative emotion induces an activated spectral profile in subcortical areas (more fast activity and less slow oscillation), while positive emotion induces a similar activation in the prefrontal cortex. However, raw power can be confounded by 1/f background; for instance, a flattening of the spectrum will inherently produce what appears to be a broad reduction in low-frequency power and an elevation in broadband high-frequency power.

### Spectral power adjusted for aperiodic component

To disentangle true oscillatory changes from shifts in the aperiodic background, we examined the aperiodic-unadjusted and -adjusted band power (i.e., the power before and after removing the 1/f fit) (**Figure 6**; **Figure S2-S4**). Interestingly, some of the valence effects on band power were altered after this adjustment. In the posterior cingulate (PCC), the apparent alpha suppression during negative emotion was no longer present in the adjusted power (*p* = 0.29); that is, once the flatter 1/f slope (due to negative emotion) was accounted for, PCC’s alpha-band power did not significantly differ from baseline (**Figure S2**). This suggests that the decrease in raw alpha power in PCC was largely a byproduct of the overall shift in the spectrum’s slope rather than a localized alpha oscillation suppression.

By contrast, in the posterior insula, a decrease in beta power during negative emotion became more pronounced in the adjusted analysis. The pIns beta reduction, which was modest in the raw spectrum (*p_FWER_* = 0.062), emerged as a significant effect after removing the aperiodic component (*p_FWER_* < 0.01). This implies that in the pIns, beyond the global flattening of the spectrum, there was a specific suppression of beta oscillatory activity linked to negative valence (**Figure S3**). In the dlPFC, the adjusted power analysis revealed that the beta-band power reduction observed in our spectral analysis may be artifactual, suggesting that positive emotion’s impact on dlPFC was confined to the broadband shift (reflected in alpha and the slope) rather than a true narrow-band beta oscillation change.

In summary, adjusting for the 1/f background clarified which band-specific changes were genuinely oscillatory. The negative-valence effects on alpha in PCC were attributable to the aperiodic shift and thus are better interpreted as part of the E/I change rather than an alpha rhythm suppression. In contrast, negative-valence effects on beta in the insula were actually stronger than raw power suggested, pointing to a targeted reduction of beta oscillatory activity in that region during aversive emotional processing. The positive-valence effects in dlPFC were mainly broadband (alpha and overall slope) with no unique beta/gamma oscillation changes. These findings highlight the value of spectral parameterization; without it, one might misinterpret broad decreases in power as multiple band-specific effects, whereas in reality, a single mechanism (e.g., change in E/I balance) can explain a coordinated shift across frequencies. (Gerster et al., 2022)

### Control analyses: Sensory confounds, physiological covariates, and scalp EEG

We conducted additional analyses to ensure that the observed neural differences were truly related to internal emotional state and not confounded by external stimulus differences or global artifacts. First, we examined whether intrinsic video features could account for the spectral changes. We quantified several properties of each video clip, including luminance (average brightness) and audio amplitude (**Figure 4B-C**). Negative clips tended to have slightly lower luminance on average (many were dark or suspenseful scenes) and higher sound variability (e.g., scenes with screams or explosions) than positive clips, which were often brighter and featured laughter or music. We entered these features as covariates in the mixed-effects models (and also looked at correlation with the neural measures directly). The valence effects remained significant even after accounting for luminance and audio levels. None of these low-level features showed a consistent relationship with the 1/f slope changes. For example, high-luminance clips did not systematically yield flatter slopes, and the thalamus/insula/cingulate effects for negative valence persisted after controlling for brightness and loudness. This suggests that the emotional content, rather than basic sensory differences, was driving the neural E/I changes.

Next, we assess whether autonomic arousal might explain the valence-related spectral differences; we incorporated trial-wise heart rate change as a covariate in the mixed-effects model. The inclusion of heart rate did not attenuate the significance of the existing valence effects, suggesting that the observed neural modulation was not simply driven by general physiological arousal but reflected valence-specific emotional processing.

Finally, we examined the simultaneously recorded scalp EEG (**Figure S5**). Scalp EEG channels also showed a modest 1/f slope change with emotion, but these effects were much smaller and not significant after correction for multiple comparisons. For example, a pIns contact showed a clear slope flattening during a disturbing clip, while simultaneously recorded occipital scalp electrodes showed only a minor change. This spatial specificity reinforces that the effects we report are not due to volume-conducted artifacts or systemic physiological changes like heart rate, which can modulate low-frequency EEG.

### Supplementary analysis: Effect of epoch length on 1/f slope estimation

In a supplementary investigation, we examined how the epoch length used for spectral analysis affects the reliability of aperiodic exponent estimation. We were motivated by the fact that emotional epochs (video clips) in our study were relatively long (30-50 s), which likely provides a robust estimate of the PSD. We recomputed the aperiodic exponent from progressively shorter segments of the video periods (e.g., truncating to 0.5 s, 2 s, etc) and observed that the estimation variance increased markedly with shorter epochs. Groupwise, the variance of the exponent across trials was inflated when using shorter windows. These observations (**Figure S6**) indicate that longer data segments are important for accurate aperiodic slope estimation, likely because the 1/f characteristic is a property of the ongoing background activity that requires sufficient sampling of the signal’s temporal structure (Gerster et al., 2022). This supplementary finding supports our use of relatively long, naturalistic stimuli; the sustained nature of the emotional engagement enabled robust measurement of each region’s spectral dynamics. It also suggests caution in future studies using very brief epochs (e.g., <2 s) to compute 1/f slopes, as this may yield unreliable or noisy estimates, potentially obscuring true effects. In our case, the long epoch lengths afforded by the video paradigm enhanced the sensitivity to detect the subtle changes in the exponent with emotional state.

## 4. Discussion

In this study, we provide novel evidence that emotional valence modulates spectral properties of human brain activity, as reflected in changes in the aperiodic (asynchronous) 1/f slope of intracranial EEG spectra. Using SEEG recordings from emotion-related cortico-limbic regions, we found that negative and positive emotional states elicited opposite patterns of 1/f slope modulation in distinct neural circuits. Negative valence with viewing aversive or unpleasant videos led to a flatter 1/f slope (less negative exponent) in key subcortical regions, including the thalamus, posterior insula, and posterior cingulate, whereas positive valence with viewing pleasant videos selectively flattened the slope in the dorsolateral prefrontal cortex. A flatter slope corresponds to relatively elevated high-frequency background activity, which has been interpreted in prior work as increased cortical excitability, reduced low-frequency synchronization, or greater neural noise in local networks (Voytek et al., 2015, Gao et al., 2017, Brake et al., 2024). Thus, our findings suggest that emotional valence alters broadband spectral features of specific circuits, highlighting the sensitivity of the aperiodic slope to shifts in internal state.

A flatter slope could indicate a state of network disinhibition or increased excitation. However, in negative emotional states, one might expect that inhibitory processes (such as fear-related interneuron activity) also play a role (Singh and Topolnik, 2023). It is possible that the flattening observed in subcortical regions during negative emotion reflects a complex interplay between excitation and inhibition: increased firing of inhibitory interneurons can paradoxically boost highfrequency LFP power, since these interneurons often fire at gamma frequencies (Tukker et al., 2007), thereby reducing low-frequency coherence and flattening the PSD. In other words, negative valence may recruit inhibitory circuitry in subcortical and midline areas as a regulatory response, but the net effect is an increase in high-frequency activity produced either directly by the inhibitory neurons themselves or indirectly by the resulting disinhibition of local pyramidal cells (Tukker et al., 2007). This interpretation – that negative emotion drives emotion-related subcortical-limbic circuits into a high-engagement state involving both excitation and inhibition – is speculative but aligns with models of emotion regulation in which subcortical regions show increased neural firing during negative affect, even as some of that activity is aimed at dampening emotional output.

In contrast, the flattening of the slope in dlPFC during positive valence suggests that pleasant emotional experiences engage executive or reward-related networks in a way that elevates broadband neural activity. The dlPFC is not classically a primary reward area, but it is implicated in working memory and attention to emotional stimuli, and in top-down regulation of emotion (Koenigs and Grafman, 2009). One possibility is that positive stimuli, by being rewarding or salient, trigger greater dorsolateral prefrontal engagement (e.g., maintaining context or monitoring the positive experience), thereby raising its local network activity. Alternatively, some of our positive clips involved humor or social content that demands cognitive appraisal, which could preferentially activate the lateral PFC. The net effect is a local shift in PFC circuit dynamics that favors higher-frequency (excitation-dominated) activity during positive emotion, hence flattening the slope. Overall, these findings support the notion that emotional valence differentially engages neural circuits – negative emotions strongly activate core subcortical areas with rapid shifts in their activity state, whereas positive emotions recruit more lateral prefrontal circuits associated with motivation and regulation.

It is important to note that not all emotion-related cortico-limbic regions responded identically to negative valence. For example, we did not observe a significant 1/f slope change in the amygdala, a typical hub for negative affect, especially fear. This may reflect the amygdala’s preferential engagement in phasic, event-related responses to salient treats rather than in the sustained background activity captured by our epoch-level analysis (Somerville et al., 2010) (**Figure S7**). Indeed, the negative-valence videos in our paradigm were not designed to elicit acute fear; rather, they involved a calm news broadcast describing distressing events. Even the video that participants rated as most negative – a new report about murder – did not include graphic or provocative imagery that would typically evoke an immediate fear response. The posterior insula and posterior cingulate, by contrast, may reflect more sustained aspects of the aversive state, such as the continuous feeling of discomfort or negative self-referential processing. The heterogeneity of slope responses across different subcortical nodes likely reflects the complex architecture of negative emotional states. For instance, a fearful stimulus might simultaneously activate circuits for sensory threat processing (insula) (Preuschoff et al., 2008), autonomic response (hypothalamus, not recorded here) (Battaglia and Thayer, 2022), subjective feeling and memory (amygdala, hippocampus) (LeDoux and Hofmann, 2018), and appraisal/self-monitoring (cingulate) (Livneh et al., 2012). Each of these subcircuits may undergo different spectral adjustments. Our results spotlight a subset (pIns, PCC, thalamus) that showed a clear spectral tilt with negative valence. It would be interesting in future studies to parse negative emotions into subtypes (fear vs. disgust vs. sadness) to see if the spectral slope changes differ across regions. For instance, the insula might be more engaged during disgust or pain, whereas the posterior cingulate might change during sadness. The current findings lay the groundwork by showing proof-of-concept that negative affect modulates local spectral slope, while also prompting deeper investigation into which emotional subprocesses drive each region’s response.

Methodologically, our study demonstrates the value of the 1/f slope as a viable neural marker for affective state. Traditional emotion electrophysiology has often focused on evoked potentials or changes in specific oscillations like frontal alpha asymmetry in EEG (de Aguiar Neto and Rosa, 2019). Here we show that parameterizing the aperiodic background provides a complementary view; the brain’s emotive state involves global changes that are detectable as slope shifts. We leveraged the FOOOF algorithm to dissociate these shifts from genuine oscillatory changes, thereby avoiding misinterpretation of broadband power increases (Donoghue et al., 2020). This approach proved critical in revealing, for example, that the PCC’s alpha-band decrease was not a true narrow-band suppression but part of a broadband change. It validates the idea that spectral slope can track internal state changes even in the absence of a prominent event-related potential or oscillatory modulation. Moreover, the consistency of slope changes within specific regions across trials suggests the measure is reliable and sensitive. We also underscore the importance of sufficient data length for slope estimation as an insight that methodological papers have noted (Gerster et al., 2022) and which we confirmed empirically. By using naturalistic stimuli that sustained an emotion over ten seconds, we obtained stable slope readings, whereas a short burst of emotion might have been harder to quantify in this manner. Thus, beyond the neuroscience findings, our study provides a template for using intracranial recordings and spectral parameterization to investigate subtle changes in brain state. This could be extended to other cognitive or clinical domains where internal states fluctuate.

The finding that the 1/f slope tracks emotional valence in specific brain circuits raises the tantalizing possibility of developing biomarkers for mood disorders based on this measure. For instance, one could hypothesize that patients with major depression might show an abnormally steep or flat slope in emotion-related cortico-limbic area at baseline, reflecting their altered baseline state of those circuits (e.g., a shift in their typical excitation/inhibition balance). In fact, recent work has hinted that the aperiodic exponent of intracranial EEG in patients with treatment-resistant depression correlates with depression severity (Hacker et al., 2025) – when patients feel more depressed, their frontal lobe 1/f slope becomes steeper (less high-frequency activity), which might relate to hypoactivation. Conversely, mania or states of hyperarousal could manifest as a very flat slope (excess high-frequency noise). Our results suggest that even in healthy individuals, transient valence shifts can push circuits toward a more extreme state of activation (either heightened or reduced activity). If a patient’s brain state is persistently in one of these extremes (e.g., subcortical regions chronically hyper-excitable in anxiety, or frontal regions chronically under-active in depression), the 1/f measure might reflect this.

Additionally, interventions for mood disorders, whether pharmacological (e.g., SSRIs, ketamine) or neuromodulatory (e.g., transcranial magnetic stimulation), could be assessed by their impact on the 1/f slope in relevant circuits. For example, ketamine, which is used for refractory depression, is known to acutely flatten the EEG power spectrum slope (De la Salle et al., 2016, Campbell et al., 2025), an effect that aligns with its transient glutamatergic surge; intriguingly, that acute effect might transiently normalize an otherwise steep slope in depressed brains, potentially correlating with symptom relief. While much more research is needed, our study positions the 1/f slope as a candidate biomarker for the state of emotional brain circuits, offering a quantitative index that could be tracked over time or used to evaluate treatment response in disorders of mood and affect.

An advantage of our study was the use of naturalistic, film-based stimuli to evoke emotions, which likely engages the brain in a way that closely resembles real-life emotional experiences, unlike brief static images or words often used in labs. This ecological validity comes with tradeoffs: the stimuli are more complex and variable, which we addressed through careful controls for content features. The fact that robust neural differences emerged despite the variability attests to the strength of the emotional manipulation. We found that longer stimulus epochs not only improved our measurement of spectral features but may also be necessary to engage a complete emotional response. A short, 1-2 s fearful image might evoke a transient amygdala potential, but a 30 s suspenseful scene can induce a sustained state of apprehension that involves widespread network changes, such as those we measured in the insula, thalamus, and cingulate (Somerville et al., 2010). Therefore, for studies aiming to probe the spectral 1/f dynamics of emotion, using protracted, immersive stimuli may be a beneficial strategy for tracking emotion on longer timescales.

On the technical side, our analysis suggests that one should ensure a baseline period is included to isolate true changes, since we saw differences relative to baseline rather than absolute power value. It also highlights that when comparing conditions that might differ in arousal or engagement, controlling for confounds is important – we took pains to show that low-level differences did not drive the effects. Moreover, when we incorporated heart rate as a physiological covariate to account for autonomic arousal, the valence effects remained robust, indicating that the neural modulations were not simply driven by global arousal but reflected emotion-specific processes. Future work could extend this approach by integrating additional physiological measures, such as skin conductance, to further map the coupling between autonomic and neural markers of emotion, thereby refining our understanding of how arousal and valence jointly shape cortical excitation–inhibition dynamics (Borah et al., 2024).

Our findings on emotional valence can be framed in the broader context of how the 1/f slope indexes brain states. Multiple studies have observed a pattern: heightened arousal or neural activation tends to flatten the slope, whereas reduced arousal or strong global inhibition tends to steepen it (Muthukumaraswamy and Liley, 2018, Colombo et al., 2019). Negative emotional states – especially high-arousal negative states like fear or stress – fit into this pattern as states of high central arousal, consistent with our observation of slope flattening in arousal-related limbic regions. Indeed, the changes we see in an aversive context parallel those seen in acute stress or anxiety models, showing increased cortical activation and reduced coherence. On the other end, states like drowsiness or sedation involve increased alpha/delta oscillations and steeper slopes (Manyukhina et al., 2024), essentially the opposite of what we saw during active emotion processing.

Even certain trait conditions mirror this dichotomy. For example, attention deficit hyperactivity disorder (ADHD) is associated with a flatter resting EEG slope (Ostlund et al., 2021), which has been interpreted as a signature of cortical hyper-excitability or reduced regulatory inhibition in children with attentional problems. This is intriguingly parallel to our state-related findings – during negative emotion, vigilance and central excitability are high, and we see a transient ADHD-like flattening in specific circuits. Likewise, pharmacological influences echo this scheme. Ketamine and some psychedelic states also flatten the PSD slope (increasing high-frequency power)(De la Salle et al., 2016, Campbell et al., 2025), consistent with a surge in excitatory synaptic activity or reduced inhibitory gating. In contrast, anesthetic agents like propofol or methylphenidate (MPH) cause a steep slope as the cortex enters an inhibited, highly synchronous state (Pertermann et al., 2019). By comparing our emotion-driven changes to these other conditions, we reinforce the idea that the 1/f slope is a broadly applicable index of neural activation level, albeit one agnostic to the precise underlying mechanism. Emotional valence shifts can thus be viewed as modulating arousal in specific circuits: negative emotions likely engage neuromodulatory systems (like norepinephrine, acetylcholine) that globally increase cortical gain and excitability (Medel et al., 2023), thereby flattening the spectrum in regions handling the emotional content. In contrast, positive emotions might engage dopaminergic or other reward-related neuromodulation that preferentially boosts activity in frontal networks. It would be fascinating in future work to measure neuromodulator levels in tandem with electrophysiology to directly link these chemical arousal signals to spectral slope change.

Despite the clear findings, our study has several limitations. First, the participant sample consisted of epilepsy patients, who are not entirely neurologically typical. We mitigate this by avoiding performing our experiment after any recently occurred seizure, but chronic epilepsy or medications could also subtly affect neural oscillations and 1/f characteristics. This issue is somewhat mitigated, as epilepsy patients are typically off their anti-seizure medication for the duration of the intracranial monitoring, and at the time of performing this experiment, any residual effect of their anti-seizure medication has completely worn off. Notably, analogous results have been observed in patients with depression but not epilepsy (Hacker et al., 2025). Nevertheless, caution is warranted when generalizing these absolute exponent values to the general population. Second, SEEG coverage is inherently limited to the regions where electrodes are implanted for clinical reasons, and we restricted our analyses to regions of particular interest and sampling rather than examining the entire brain. We had good coverage of some emotion-related cortico-limbic network, but our sampling may have missed effects in regions not well covered (for example, the basal ganglia might be relevant to emotion valence but were not sampled sufficiently in our cohort). Thus, the absence of evidence in a region, such as ACC, is not evidence of the absence of effect.

Third, while we discuss the aperiodic (asynchronous) slope in the context of excitation/inhibition balance – supported by some prior findings linking slope changes to cortical disinhibition or excitation (Gao et al., 2017, Medel et al., 2023) – this interpretation is speculative and indirect. Multiple mechanistic accounts of aperiodic activity have been proposed. For example, the intrinsic low-pass filtering properties of dentrites can contribute to 1/f spectral scaling independent of spiking dynamics (Buzsáki et al., 2012), and the frequency-dependent conductance of extracellular space (e.g., due to ionic diffusion) can also produce 1/f-like spectra (Bédard and Destexhe, 2009). Moreover, a superposition of stochastically driven oscillatory processes with different relaxation rates can mathematically yield a 1/f slope without invoking any changes in synaptic excitation or inhibition (Evertz et al., 2022). Aperiodic activity may thus reflect a more global measure of neuronal noise or network complexity (Voytek et al., 2015, Kramer and Chu, 2024), rather than a specific ratio of excitatory to inhibitory currents. In fact, recent experimental work has questioned the aperiodic exponent’s reliability as a marker of E/I balance. For instance, selectively suppressing interneuron activity in the mouse cortex paradoxically increased the exponent instead of flattening it (Salvatore et al., 2024). Thus, we avoid attributing our effects to any single microcircuit mechanism. While our results are consistent with the notion of valence-related shifts in cortical excitability, establishing a direct causal link between spectral slope changes and a specific E/I shift would require concurrent measurement of underlying inhibitory and excitatory activity. Animal studies or computational models will be needed to validate whether the valence-driven slope changes observed here indeed correspond to specific changes in excitation/inhibition balance, or are better explained by other physiological processes.

Finally, the emotional stimuli, while powerful, were not uniform in content, and emotional experiences are subjective. Some variability in neural response likely comes from the differences in how each person internally reacted to a given video (e.g., one person’s fear might be another’s disgust). Our sample size (n = 31), while typical for human subject intracranial studies, limits statistical power to detect more minor effects and to explore finer-grained emotional distinctions. Despite these limitations, the consistency of specific region-specific effects across individuals gives confidence in the robustness of the core findings.

In conclusion, this work demonstrates that the aperiodic 1/f slope of the human intracranial EEG is sensitive to the valence of emotional experiences. Positive and negative emotions drive distinct shifts in this spectral measure in prefrontal versus subcortical circuits, respectively, reflecting changes in the local neural population dynamics. These findings bridge a gap between animal studies of microcircuit physiology and human affective neuroscience by providing a window into how the human brain’s rhythmic and arrhythmic activity dynamically reconfigure with subjective emotional state. The 1/f slope emerges as a promising quantitative biomarker of internal brain states – here validated in the context of emotion – and future research may establish its utility in monitoring and modulating mood in clinical settings. As we refine our understanding of this measure, it could aid in the development of new therapies for mood and anxiety disorders, guiding interventions that seek to restore healthy neural balance in dysregulated neural circuits. Ultimately, by quantifying these fundamental aspects of neural circuit function, we move one step closer to a physiologically informed understanding of emotion, wherein metrics like the spectral slope can index the otherwise invisible neural changes that underlie the ebb and flow of mood.

## Acknowledgments

We express our gratitude to all the patients who participated in this study; to the neurologists, nurses, and technicians in the Epilepsy Monitoring Unit at Barnes-Jewish Hospital who helped make this research possible. This work was supported by the NIH/NIBIB (P41-EB018783, R01-EB026439), NIH/NINDS (U24-NS109103, U01-NS108916, U01-NS128612, R21-NS128307), NIH/NIMH (R01-MH120194, R01-MH122258), McDonnell Center for Systems Neuroscience, and Fondazione Neurone.

## Authors’ Contributions

Conceptualization: C.H.; Methodology: C.H.; Software: H.C., P.B. and H.P.; Validation: H.C., T.X. and A.S.; Formal Analysis: H.C. and H.P.; Investigation: H.C., G.T., A.S., H.P., P.B. and J.T.W.; Resources: P.B. and J.T.W.; Data Curation: P.B.; Writing-Original Draft: H.P.; Writing-Review and Editing: H.P., P.B. and J.T.W.; Visualization: H.C., G.T. and H.P.; Supervision: P.B. and J.T.W; Project Administration: P.B. and J.T.W.; Funding Acquisition: P.B. and J.T.W..

All authors read and approved the final version of the manuscript.

## Competing Interests

The authors declare that we have no competing interests.

## Data Availability

Comprehensive data-sets may be provided to interested researchers upon reasonable request to the corresponding author and approval of a data-sharing agreement by the Institutional Review Board of Washington University in St. Louis.

## Code Availability

The MATLAB scripts and the data necessary to reproduce the results presented in this manuscript will be made available without restrictions on the National Center for Adaptive Neurotechnologies GitHub repository.

## Declaration of generative AI and AI-assisted technologies in the writing process

During the preparation of this work, the authors utilized ChatGPT and Grammarly to enhance language clarity and readability. After using these tools, the authors reviewed and edited the content as needed, taking full responsibility for the publication’s content.

## Supplementary Figures

**Table S1.** Demographical information. A total of thirty-one patients enrolled in this study. We had three left, three ambidextrous, and one mixed-handed participant. The average IQ was 99.9. All participants had an IQ higher than 71, which was our criterion for enrolling in the study. We had fourteen females in our study. The mean age was 34.6 years. On average, 204.3 electrodes were implanted per patient. The number of electrodes represents the total number of electrodes implanted, including those excluded from our analysis.

| Subject | Handedness | IQ | Sex | Age range | #Electrodes |
| --- | --- | --- | --- | --- | --- |
| S01 | Right | n/a | Male | 41-45 | 110 |
| S02 | Right | n/a | Female | 46-50 | 177 |
| S03 | Right | n/a | Male | 21-25 | 220 |
| S04 | Right | 100 | Male | 25-30 | 218 |
| S05 | Right | 95 | Male | 26-30 | 184 |
| S06 | Right | 90 | Female | 36-40 | 226 |
| S07 | Right | 80 | Male | 66-70 | 172 |
| S08 | Right | n/a | Female | 36-40 | 229 |
| S09 | Right | 98 | Male | 36-40 | 150 |
| S10 | Right | 76 | Male | 36-40 | 214 |
| S11 | Right | n/a | Female | 36-40 | 229 |
| S12 | Right | 107 | Female | 31-35 | 190 |
| S13 | Right | 88 | Female | 21-25 | 222 |
| S14 | Right | n/a | Male | 26-30 | 266 |
| S15 | Right | 104 | Female | 41-45 | 202 |
| S16 | Ambidextrous | 106 | Male | 41-45 | 162 |
| S17 | Ambidextrous | 105 | Male | 21-25 | 226 |
| S18 | Left | 89 | Female | 56-60 | 229 |
| S19 | Right | 108 | Female | 36-40 | 182 |
| S20 | Right | 111 | Male | 21-25 | 274 |
| S21 | Right | 85 | Male | 36-40 | 219 |
| S22 | Right | 111 | Female | 36-40 | 229 |
| S23 | Ambidextrous | 105 | Male | 21-25 | 229 |
| S24 | Left | 103 | Female | 51-55 | 211 |
| S25 | Right | 111 | Male | 26-30 | 223 |
| S26 | Left | 71 | Male | 36-40 | 229 |
| S27 | Mixed | 120 | Male | 26-30 | 204 |
| S28 | Right | 117 | Female | 26-30 | 142 |
| S29 | Right | 105 | Male | 21-25 | 186 |
| S30 | Right | 110 | Female | 21-25 | 254 |
| S31 | Right | 102 | Female | 16-20 | 126 |
| Average | 3 Left / 24 Right | 100 | 17 M /14 F | 35 | 204 |

| Subject | Handedness | IQ | Sex | Age range | #Electrodes |
| --- | --- | --- | --- | --- | --- |

**Table S2.** Number of electrodes distributed across thalamic nucleus. Of 180 thalamic electrodes, 131 were assigned to a specific thalamic nucleus. Nuclei were grouped into five categories: anterior (ANT), medial (MNT), ventral (VNT), and posterior (PNT) nuclei of the thalamus, and interlaminar thalamic nuclei (ITN). ANT comprised the anteroventral nucleus (AV). ITN includes the central medial (CeM), central lateral (CL), centromedian (CM) nucleus, and parafascicular (Pf). MNT includes the medial dorsal (MD) and medial ventral (MV) nucleus. VNT includes the ventral anterior (VA), ventral lateral (VL), ventral posterior (VP), and ventral medial (VM) nucleus. PNT includes pulvinar nuclei and geniculate bodies. The mismatch between the total number of thalamic electrodes (**Figure 2B**) and the sum of labeled nuclei electrodes is because some of the electrodes that could not be assigned to any of the nuclei listed above.

| Thalamic nucleus | #Electrodes | #Subjects |
| --- | --- | --- |
| ANT (AV) | 11 | 11 |
| ITN | 18 | 14 |
| CeM | 4 | 3 |
| CL | 2 | 1 |
| CM | 11 | 9 |
| Pf | 1 | 1 |
| MNT | 6 | 4 |
| MD | 6 | 4 |
| MV | 0 | 0 |
| VNT | 73 | 19 |
| VA | 28 | 12 |
| VL | 33 | 12 |
| VP | 11 | 5 |
| VM | 1 | 1 |
| PNT | 23 | 7 |
| Pulvinar | 23 | 7 |
| Geniculate | 0 | 0 |

**Figure S1.**
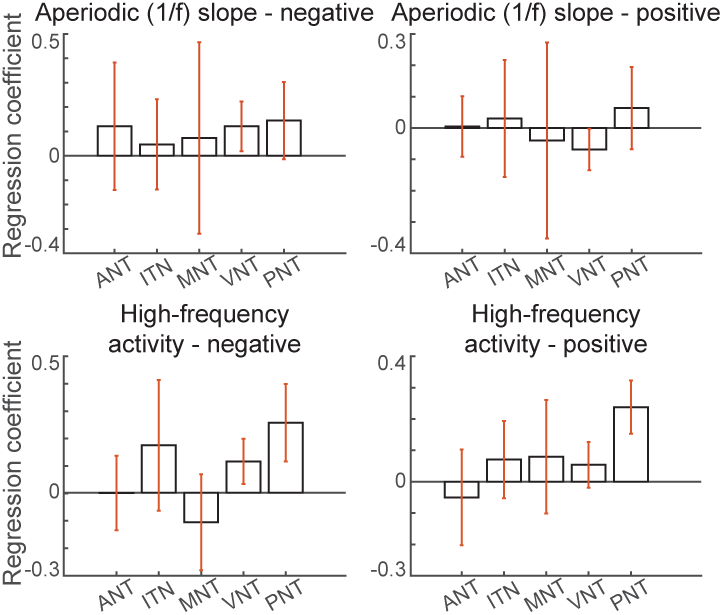
Aperiodic slope and high-frequency activity in thalamic nuclei. During viewing of negative video stimuli, the aperiodic slope flattened (i.e., increased) in ventral nuclei (VNT; top left), and high-frequency activity increased in both the VNT and posterior nuclei (PNT; bottom left). In contrast, during viewing of positive video stimuli, no nucleus showed a significant change in aperiodic slope (top right), whereas high-frequency activity increased in the PNT. Notably, it is worth taking attention to the concurrent increment of the aperiodic exponent and high-frequency power mirrors the pattern observed in the whole-thalamus analysis (**Figure 5**). Given the pulvinar’s established role, which represents the visual attention and salience (Zhou et al., 2016), the high-frequency activity increment of the pulvinar in both negative and positive stimuli may be reflected by these functions. Error bars (orange) indicates 99% confidence intervals.

**Figure S2.**
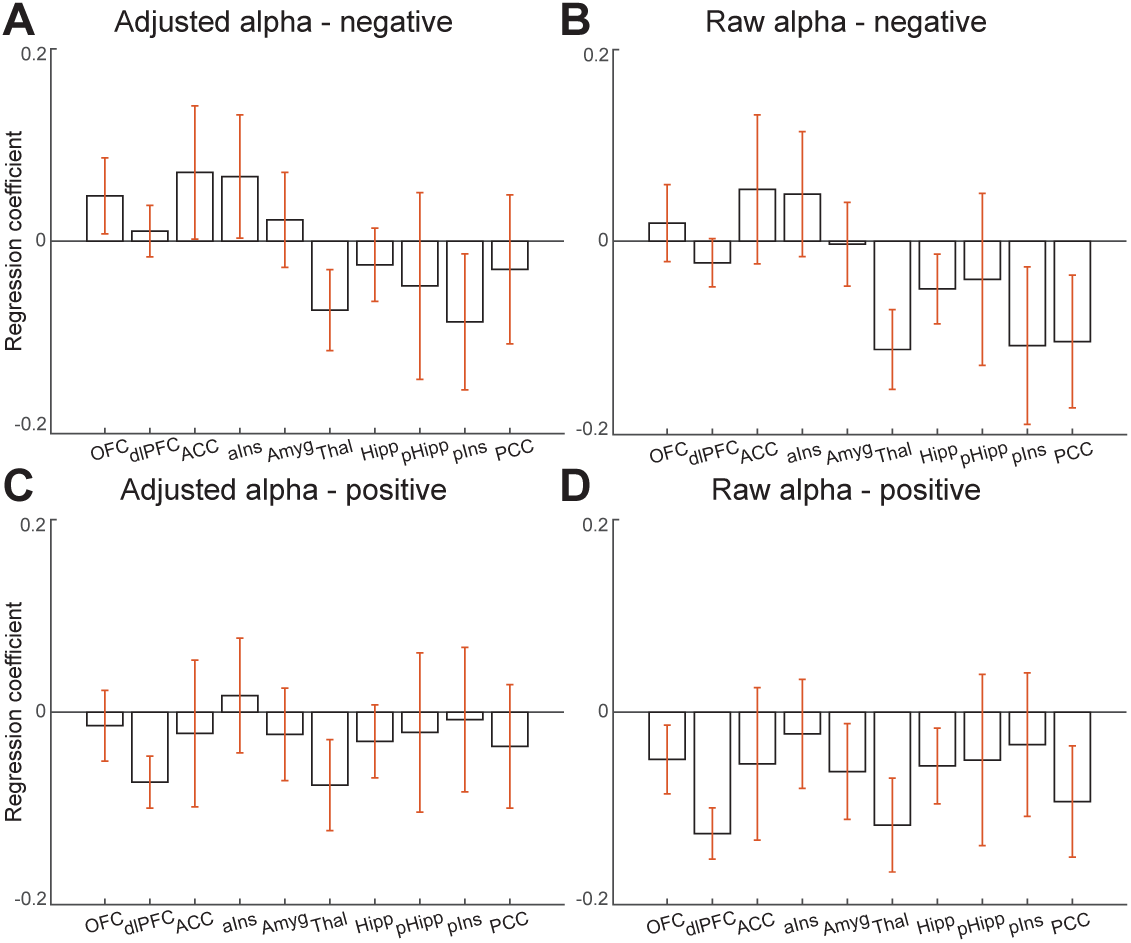
Alpha power before and after 1/f-adjustment for all regions. **(A-B)** Thalamic and posterior insula alpha activity has decreased during viewing the negative emotional video. OFC, ACC, and anterior insular alpha power increased during viewing a negative emotional video, while it gained significance after 1/f-adjustment. PCC alpha activity reduction lost its significance after 1/f-adjustment. **(C-D)** Dorsolateral PFC, thalamic alpha power has decreased during viewing the positive emotional video. OFC, amygdalar, hippocampal, and PCC alpha power reduction has lost its significance after 1/f-adjustment.

**Figure S3.**
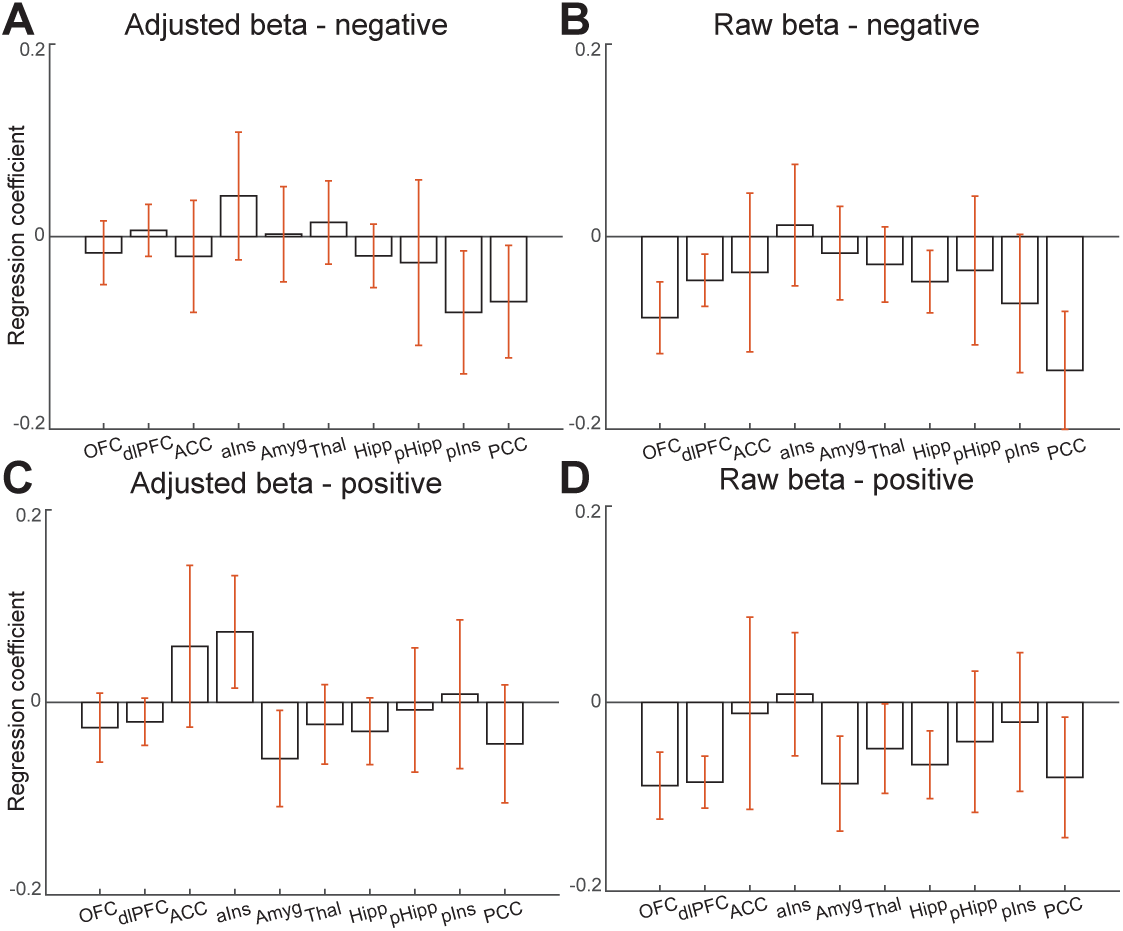
Beta power before and after 1/f-adjustment for all regions. **(A-B)** PCC beta activity has decreased while watching the negative emotional video. Posterior insular beta activity has decreased during viewing a negative emotional video, while it gained significance after 1/f-adjustment. OFC, dorsolateral PFC, and hippocampal beta activity ended up not decreasing significantly during the negative emotional video after 1/f-adjustment. **(C-D)** Amygdalar beta activity has decreased during viewing a positive emotional video. Anterior insular beta activity has increased during viewing positive emotional video, while it gained significance after 1/f-adjustment. OFC, dorsolateral PFC, thalamic, hippocampal, and PCC beta activity reduction has lost its significance after 1/f-adjustment.

**Figure S4.**
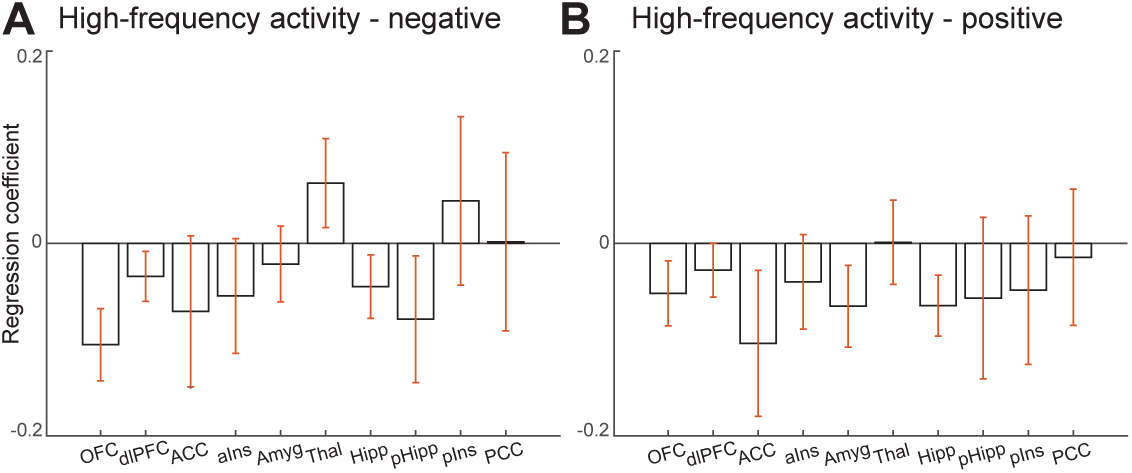
High-frequency activity (HFA) for all regions. **(A)** Thalamic HFA has increased during viewing a negative emotional video, while the OFC, dorsolateral PFC, hippocampal, and parahippocampal HFA have decreased during viewing the negative emotional videos. **(B)** OFC, ACC, amygdala, and hippocampal HFA have decreased during viewing the positive emotional videos. The orange error bar indicates a 99.5% confidence interval.

**Figure S5.**
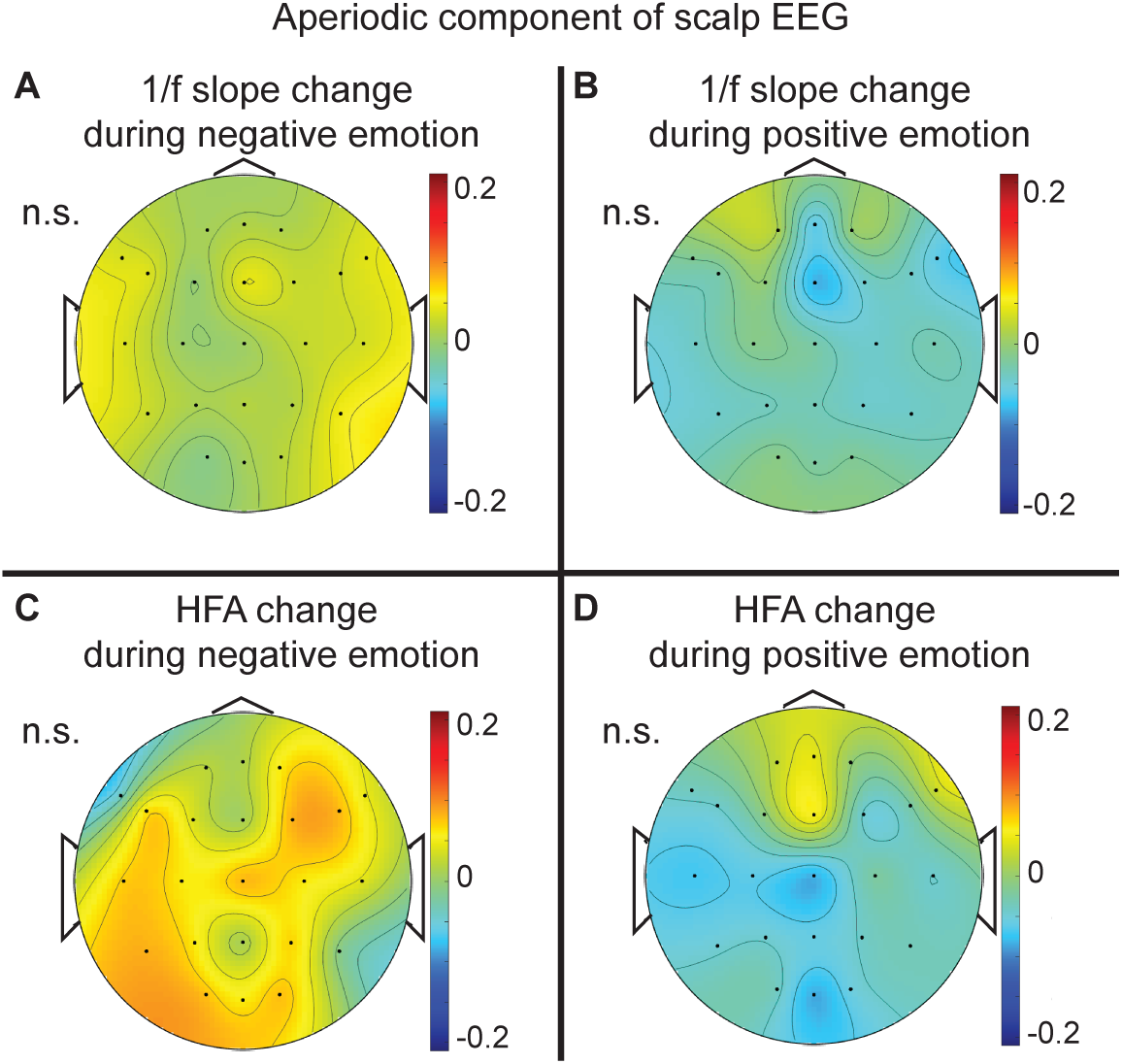
Scalp EEG results. None of the scalp electrodes showed significant 1/f and HFA changes during both positive and negative video.

**Figure S6.**
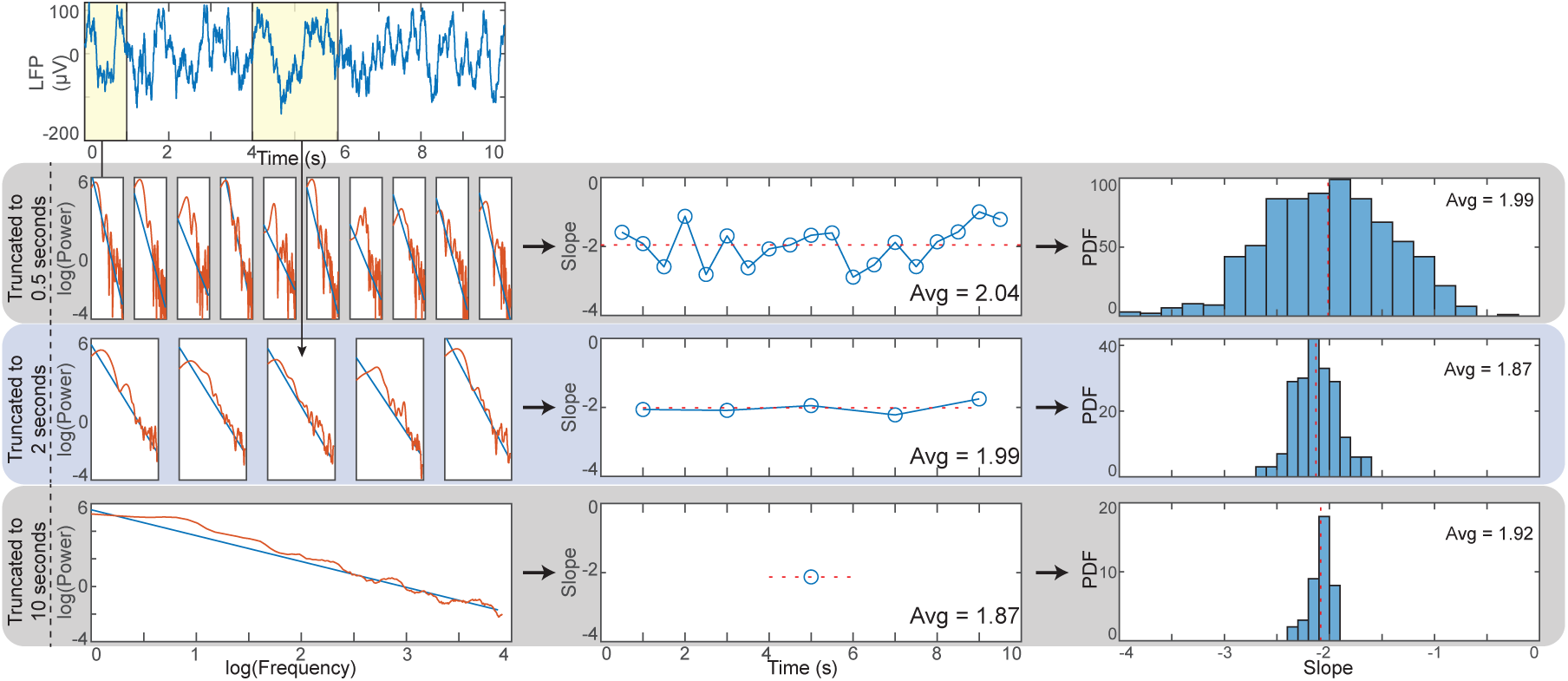
Epoch length affects 1/f estimate variability. We used different epoching lengths to estimate aperiodic slope, where they are truncated into 2s, 5s, 10s. A shorter epoch increased the variability of the 1/f slope estimate, while the average value did not change significantly.

**Figure S7.**
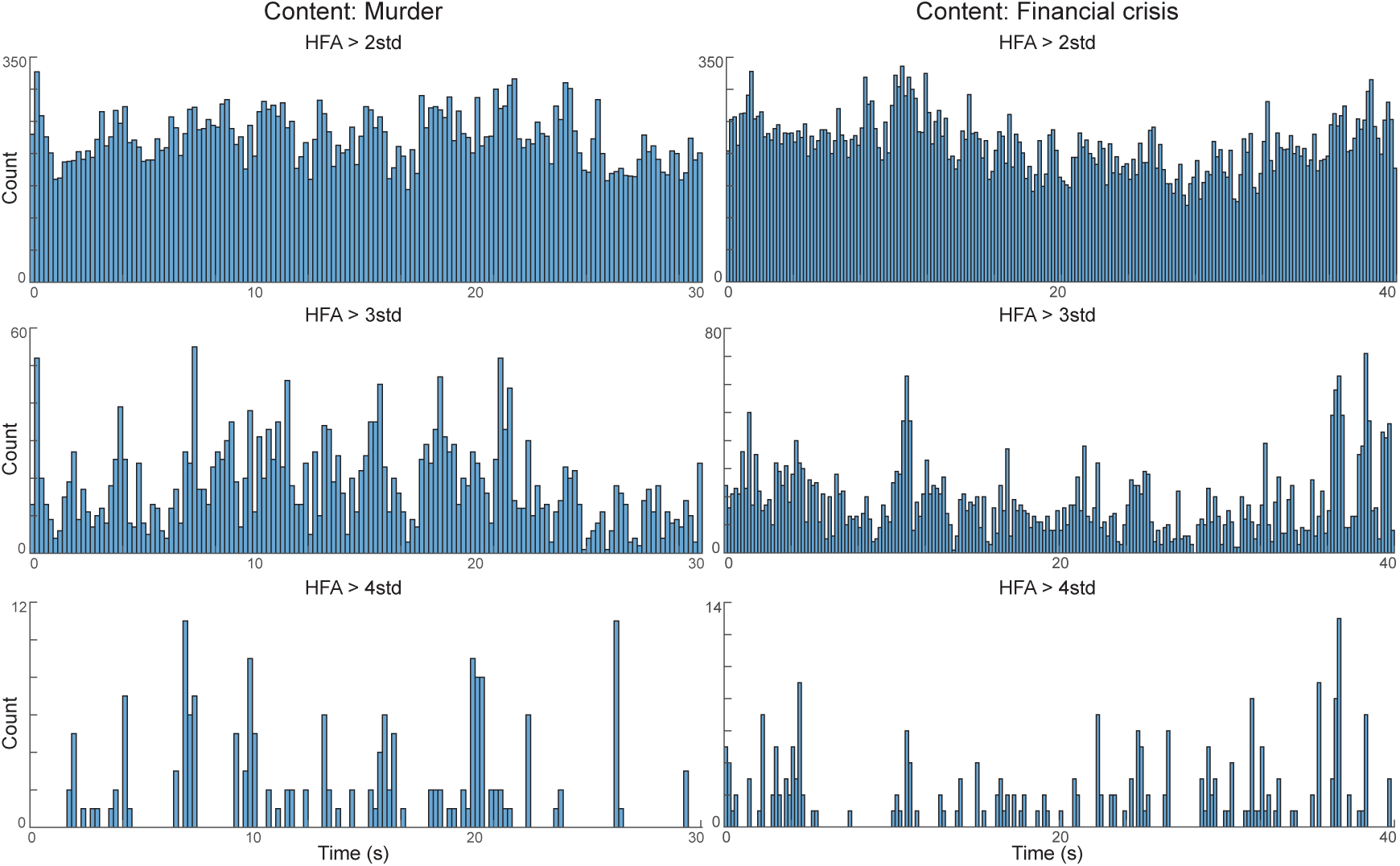
High-frequency activity of amygdala during viewing negative emotional video. High-frequency activity (HFA) in the amygdala during the two videos that participants rated as the most negative – news clips about murder and financial crisis. The videos were 30 s (murder) and 40 s (financial crisis) in duration. HFA events were counted within 100 ms windows whenever the signal exceeded a predefined threshold across a total of 161 amygdala electrodes. Distributions were examined across three thresholds (2, 3, and 4 standard deviations). The analyses revealed no consistent differences in HFA between the early and late segments of the videos, nor any time points with unusually high activity relative to the number of amygdala electrodes analyzed.

## Notes

### Competing Interest Statement

The authors have declared no competing interest.

### Author Declarations

IRB of Washington University in St. Louis gave ethical approval for this work (IRB ID#: 202104033-1001).

## References

Adamek, M., Swift, J., Brunner, P., 2022. VERA - A versatile electrode localization framework, Version 1.0.0. Zenodo..

de Aguiar Neto, F.S., Rosa, J.L.G., 2019. Depression biomarkers using non-invasive EEG: A review. Neuroscience & Biobehavioral Reviews 105, 83–93.

Ahmad, J., Ellis, C., Leech, R., Voytek, B., Garces, P., Jones, E., Buitelaar, J., Loth, E., Dos Santos, F.P., Amil, A.F., et al., 2022. From mechanisms to markers: novel noninvasive EEG proxy markers of the neural excitation and inhibition system in humans. Translational psychiatry 12, 467.

Battaglia, S., Thayer, J.F., 2022. Functional interplay between central and autonomic nervous systems in human fear conditioning. Trends in Neurosciences 45, 504–506.

Bédard, C., Destexhe, A., 2009. Macroscopic models of local field potentials and the apparent 1/f noise in brain activity. Biophysical journal 96, 2589–2603.

Bönstrup, M., Hagemann, J., Gerloff, C., Sauseng, P., Hummel, F.C., 2015. Alpha oscillatory correlates of motor inhibition in the aged brain. Frontiers in Aging Neuroscience 7, 193.

Borah, R.M., Pathak, A., Banerjee, A., 2024. Emotion arousal but not valence is strongly represented in aperiodic EEG activity stemming from thalamocortical interactions. bioRxiv, 2024–03.

Brake, N., Duc, F., Rokos, A., Arseneau, F., Shahiri, S., Khadra, A., Plourde, G., 2024. A neurophysiological basis for aperiodic EEG and the background spectral trend. Nature communications 15, 1514.

Buzsáki, G., Anastassiou, C.A., Koch, C., 2012. The origin of extracellular fields and currents—EEG, ECoG, LFP and spikes. Nature reviews neuroscience 13, 407–420.

Campbell, K., Neul, J.L., Lieberman, D.N., Berry-Kravis, E., Benke, T.A., Fu, C., Percy, A., Suter, B., Morris, D., Carpenter, R.L., et al., 2025. A randomized, placebo-controlled, cross-over trial of ketamine in Rett syndrome. Journal of Neurodevelopmental Disorders 17, 4.

Cheung, C., Hamilton, L.S., Johnson, K., Chang, E.F., 2016. The auditory representation of speech sounds in human motor cortex. elife 5, e12577.

Colombo, M.A., Napolitani, M., Boly, M., Gosseries, O., Casarotto, S., Rosanova, M., Brichant, J.F., Boveroux, P., Rex, S., Laureys, S., et al., 2019. The spectral exponent of the resting EEG indexes the presence of consciousness during unresponsiveness induced by propofol, xenon, and ketamine. NeuroImage 189, 631–644.

Coon, W.G., Gunduz, A., Brunner, P., Ritaccio, A.L., Pesaran, B., Schalk, G., 2016. Oscillatory phase modulates the timing of neuronal activations and resulting behavior. NeuroImage 133, 294–301.

Cross, Z.R., Gray, S.M., Dede, A.J., Rivera, Y.M., Yin, Q., Vahidi, P., Rau, E.M., Cyr, C., Holubecki, A.M., Asano, E., et al., 2025. The development of aperiodic neural activity in the human brain. Nature human behaviour, 1–16.

Donoghue, T., Haller, M., Peterson, E.J., Varma, P., Sebastian, P., Gao, R., Noto, T., Lara, A.H., Wallis, J.D., Knight, R.T., et al., 2020. Parameterizing neural power spectra into periodic and aperiodic components. Nature neuroscience 23, 1655–1665.

Evertz, R., Hicks, D.G., Liley, D.T., 2022. Alpha blocking and 1/fβ spectral scaling in resting EEG can be accounted for by a sum of damped alpha band oscillatory processes. PLoS computational biology 18, e1010012.

Fischl, B., 2012. Freesurfer. Neuroimage 62, 774–781.

Gao, R., Peterson, E.J., Voytek, B., 2017. Inferring synaptic excitation/inhibition balance from field potentials. Neuroimage 158, 70–78.

Gerster, M., Waterstraat, G., Litvak, V., Lehnertz, K., Schnitzler, A., Florin, E., Curio, G., Nikulin, V., 2022. Separating neural oscillations from aperiodic 1/f activity: challenges and recommendations. Neuroinformatics 20, 991–1012.

Greicius, M.D., Krasnow, B., Reiss, A.L., Menon, V., 2003. Functional connectivity in the resting brain: a network analysis of the default mode hypothesis. Proceedings of the national academy of sciences 100, 253–258.

Hacker, C., Mocchi, M.M., Xiao, J., Metzger, B., Adkinson, J., Pascuzzi, B., Mathura, R., Oswalt, D., Watrous, A., Bartoli, E., et al., 2025. Aperiodic (1/f) neural activity robustly tracks symptom severity changes in treatment-resistant depression. Biological Psychiatry: Cognitive Neuroscience and Neuroimaging 10, 186–194.

Hamilton, J.P., Farmer, M., Fogelman, P., Gotlib, I.H., 2015. Depressive rumination, the default-mode network, and the dark matter of clinical neuroscience. Biological psychiatry 78, 224–230.

Haroon, E., Chen, X., Li, Z., Patel, T., Woolwine, B.J., Hu, X.P., Felger, J.C., Miller, A.H., 2018. Increased inflammation and brain glutamate define a subtype of depression with decreased regional homogeneity, impaired network integrity, and anhedonia. Translational psychiatry 8, 189.

Kim, J., Kim, H., Kim, S., Park, H., Fava, M., Mischoulon, D., Jeon, H.J., Jeong, B., 2025. Impulsive loss decision-making associated with aberrant meso-/habenular-cortical functional networks in young adults with major depressive disorder with suicidal ideation. Journal of Affective Disorders, 120074.

Koenigs, M., Grafman, J., 2009. The functional neuroanatomy of depression: distinct roles for ventromedial and dorsolateral prefrontal cortex. Behavioural brain research 201, 239–243.

Kramer, M.A., Chu, C.J., 2024. A general, noise-driven mechanism for the 1/f-like behavior of neural field spectra. Neural computation 36, 1643–1668.

LeDoux, J.E., Hofmann, S.G., 2018. The subjective experience of emotion: a fearful view. Current opinion in behavioral sciences 19, 67–72.

Livneh, U., Resnik, J., Shohat, Y., Paz, R., 2012. Self-monitoring of social facial expressions in the primate amygdala and cingulate cortex. Proceedings of the National Academy of Sciences 109, 18956–18961.

Lozano-Soldevilla, D., Ter Huurne, N., Cools, R., Jensen, O., 2014. GABAergic modulation of visual gamma and alpha oscillations and its consequences for working memory performance. Current Biology 24, 2878–2887.

Manyukhina, V.O., Prokofyev, A.O., Obukhova, T.S., Stroganova, T.A., Orekhova, E.V., 2024. Changes in high-frequency aperiodic 1/f slope and periodic activity reflect post-stimulus functional inhibition in the visual cortex. Imaging Neuroscience 2, 1–24.

Medel, V., Irani, M., Crossley, N., Ossandón, T., Boncompte, G., 2023. Complexity and 1/f slope jointly reflect brain states. Scientific reports 13, 21700.

Miller, K.J., Sorensen, L.B., Ojemann, J.G., Den Nijs, M., 2009. Power-law scaling in the brain surface electric potential. PLoS computational biology 5, e1000609.

Muthukumaraswamy, S.D., Liley, D.T., 2018. 1/f electrophysiological spectra in resting and drug-induced states can be explained by the dynamics of multiple oscillatory relaxation processes. NeuroImage 179, 582–595.

Ostlund, B.D., Alperin, B.R., Drew, T., Karalunas, S.L., 2021. Behavioral and cognitive correlates of the aperiodic (1/f-like) exponent of the EEG power spectrum in adolescents with and without ADHD. Developmental cognitive neuroscience 48, 100931.

Park, H., Kim, M., Kim, J., Kim, S., Jeong, B., 2025. Anterior insular cortex glutamate-glutamine (Glx) levels predict general psychopathology via heightened error sensitivity. Frontiers in Neuroscience 19, 1592015.

Pertermann, M., Bluschke, A., Roessner, V., Beste, C., 2019. The modulation of neural noise underlies the effectiveness of methylphenidate treatment in attention-deficit/hyperactivity disorder. Biological Psychiatry: Cognitive Neuroscience and Neuroimaging 4, 743–750.

Preuschoff, K., Quartz, S.R., Bossaerts, P., 2008. Human insula activation reflects risk prediction errors as well as risk. Journal of Neuroscience 28, 2745–2752.

Quon, R.J., Meisenhelter, S., Camp, E.J., Testorf, M.E., Song, Y., Song, Q., Culler, G.W., Moein, P., Jobst, B.C., 2022. AiED: Artificial intelligence for the detection of intracranial interictal epileptiform discharges. Clinical neurophysiology 133, 1–8.

De la Salle, S., Choueiry, J., Shah, D., Bowers, H., McIntosh, J., Ilivitsky, V., Knott, V., 2016. Effects of ketamine on resting-state EEG activity and their relationship to perceptual/dissociative symptoms in healthy humans. Frontiers in Pharmacology 7, 348.

Salvatore, S.V., Lambert, P.M., Benz, A., Rensing, N.R., Wong, M., Zorumski, C.F., Mennerick, S., 2024. Periodic and aperiodic changes to cortical EEG in response to pharmacological manipulation. Journal of neurophysiology 131, 529–540.

Samide, R., Cooper, R.A., Ritchey, M., 2020. A database of news videos for investigating the dynamics of emotion and memory. Behavior Research Methods 52, 1469–1479.

Sarawagi, A., Soni, N.D., Patel, A.B., 2021. Glutamate and GABA homeostasis and neurometabolism in major depressive disorder. Frontiers in psychiatry 12, 637863.

Satterthwaite, F.E., 1946. An approximate distribution of estimates of variance components. Biometrics bulletin 2, 110–114.

Schalk, G., McFarland, D.J., Hinterberger, T., Birbaumer, N., Wolpaw, J.R., 2004. BCI2000: a general-purpose brain-computer interface (BCI) system. IEEE Transactions on biomedical engineering 51, 1034–1043.

Singh, S., Topolnik, L., 2023. Inhibitory circuits in fear memory and fear-related disorders. Frontiers in neural circuits 17, 1122314.

Somerville, L.H., Whalen, P.J., Kelley, W.M., 2010. Human bed nucleus of the stria terminalis indexes hypervigilant threat monitoring. Biological psychiatry 68, 416–424.

Tukker, J.J., Fuentealba, P., Hartwich, K., Somogyi, P., Klausberger, T., 2007. Cell type-specific tuning of hippocampal interneuron firing during gamma oscillations in vivo. Journal of Neuroscience 27, 8184–8189.

Voytek, B., Kramer, M.A., Case, J., Lepage, K.Q., Tempesta, Z.R., Knight, R.T., Gazzaley, A., 2015. Age-related changes in 1/f neural electrophysiological noise. Journal of neuroscience 35, 13257–13265.

Williams, L.M., 2016. Precision psychiatry: a neural circuit taxonomy for depression and anxiety. The Lancet Psychiatry 3, 472–480.

Xiao, J., Provenza, N.R., Asfouri, J., Myers, J., Mathura, R.K., Metzger, B., Adkinson, J.A., Allawala, A.B., Pirtle, V., Oswalt, D., et al., 2023. Decoding depression severity from intracranial neural activity. Biological psychiatry 94, 445–453.

Zhou, H., Schafer, R.J., Desimone, R., 2016. Pulvinar-cortex interactions in vision and attention. Neuron 89, 209–220.

